# The genetic architecture of human infectious diseases and pathogen-induced cellular phenotypes

**DOI:** 10.1101/2020.07.19.20157404

**Authors:** Andrew T. Hale, Dan Zhou, Rebecca L. Sale, Lisa Bastarache, Liuyang Wang, Sandra S. Zinkel, Steven J. Schiff, Dennis C. Ko, Eric R. Gamazon

## Abstract

Here, we develop a genetics-anchored framework to decipher mechanisms of infectious disease (ID) risk and infer causal effect on potential complications. We perform transcriptome-wide association studies (TWAS) of 35 ID traits in 27,615 individuals in a broad collection of human tissues, identifying 70 gene-level associations with 26 ID traits, with replication in two large-scale biobanks. A phenome-scale scan and Mendelian Randomization of the 70 gene-level associations across 197 traits proposes a molecular basis for known complications of the ID traits. This rich resource of host genetic associations with pathogen cultures and 16S-rRNA-based microbiome variation provides a platform to investigate host-pathogen interactions. To identify relevant cellular processes, we develop a TWAS repository of 79 pathogen-exposure induced cellular phenotypes. Our study will facilitate mechanistic insights into the role of host genetic variation on ID risk and pathophysiology, with important implications for our molecular understanding of severe phenotypic outcomes.

## INTRODUCTION

Genome-wide association studies (GWAS) and large-scale DNA biobanks with phenome-scale information are making it possible to identify the genetic basis of a wide range of complex traits in humans^1,2^. A parallel development is the increasing availability of GWAS summary statistics, facilitating genetic analyses of entire disease classes and promising considerably improved resolution of genetic effects on human disease^3,4^. Recent analysis involving 558 well-powered GWAS results found that trait-associated loci cover ∼50% of the genome, enriched in both coding and regulatory regions, and of these, ∼90% are implicated in multiple traits^5^. However, the breadth of clinical and biological information in these datasets will require new methodologies and additional high-dimensional data to advance our understanding of the genetic architecture of complex traits and relevant molecular mechanisms^6-8^. Approaches to understanding the functional consequences of implicated loci and genes are needed to determine causal pathways and potential mechanisms for pharmacological intervention.

The genetic basis of infectious disease (ID) risk and severity has been relatively understudied using GWAS methodologies. ID risk and pathogenesis is likely to be multifactorial, resulting from a complex interplay of host genetic variation, environmental exposure, and pathogen-specific molecular mechanisms. Although monogenic mechanisms of ID risk have been demonstrated, characterizing the genetic architecture of ID risk remains challenging^9-13^. Phenome-scale information increasingly available in large-scale DNA biobanks offers opportunities to fill gaps in our understanding of the causal effect of ID risk on other traits, including adverse outcomes.

Here we conduct genome-wide association studies (GWAS) and transcriptome-wide association studies (TWAS) of 35 ID traits. To implement the latter, we apply PrediXcan^7,14^, which exploits the genetic component of gene expression to probe the molecular basis of disease risk. We combine information across a broad collection of tissues to determine gene-level associations using a multi-tissue approach, which displays improved statistical power over a single-tissue approach^7,14,15^. Notably, we identify 70 gene-level associations for 26 of 35 ID traits, i.e., heretofore referred to as ID-associated genes, and conduct replication using the corresponding traits in the UK Biobank and FinnGen consortia data^1,16^. The rich resource of genetic information linked to clinical microbiology information (serology and culture data) across bacterial, fungal, and viral genera that we leverage provides a platform to interrogate the genetic basis of compartment-specific infection and colonization. Linking high-resolution taxonomic classification from 16S ribosomal RNA (rRNA) sequencing to host GWAS information has been used to investigate the contribution of host genetic variation to microbiome composition^17^. We exploit host GWAS for 155 pathogens in the microbiome^17^ to gain further insights into identified genetic risk factors for an ID trait. To determine the phenotypic consequences of ID-associated genes, including adverse outcomes and complications, we perform a phenome-scale scan across hematologic, respiratory, cardiovascular, and neurologic traits. We use a Mendelian Randomization framework^18^ to conduct causal inference on the effect of a clinical ID trait on an adverse clinical outcome. To elucidate the cellular mechanisms through which host genetic variation influences disease risk, we generate an atlas of gene-level associations with 79 pathogen-induced cellular phenotypes determined by High-throughput Human *in vitrO* Susceptibility Testing (Hi-HOST)^19^ as a discovery and replication platform. The rich genomic resource we generate and the integrative methodology we develop promise to yield discoveries on the molecular mechanisms of infection, improve our understanding of adverse outcomes and complications, and enable prioritization of new therapeutic targets.

## RESULTS

A schematic diagram illustrating our study design and the reference resource we provide can be found in Figure 1. Here we analyzed 35 clinical ID traits, 79 pathogen-exposure-induced cellular traits, and 197 (cardiovascular, hematologic, neurologic, and respiratory) traits. We performed GWAS and TWAS^14,20^ to investigate the genetic basis of the ID traits and their potential adverse outcomes and complications. We exploited serology and culture data linked to genetic information and genome-wide associations with microbiome traits to investigate compartment-specific patterns of infection. We conducted causal inference within a Mendelian Randomization framework^21^, exploiting genetic instruments for naturally “randomized controlled trials” to evaluate the causal effect of a modifiable exposure or risk factor on a clinical phenotype. We generate a rich resource for understanding the genetic and molecular basis of infection and potential adverse effects and complications.

**Figure 1.**
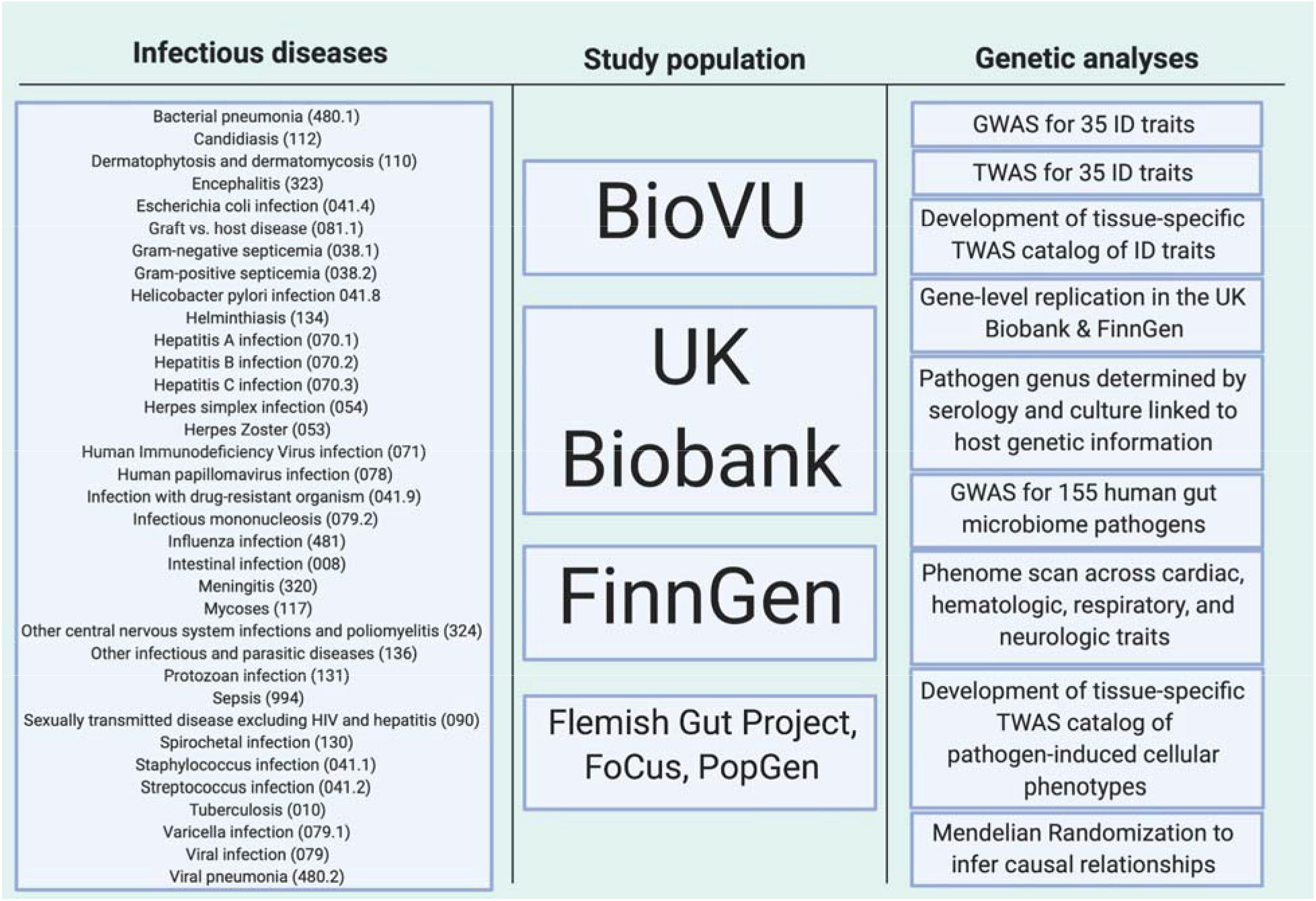
Overview of ID atlas resource. List of ID traits tested with corresponding Phecode (phewascatalog.org) in parentheses.

### GWAS and TWAS of 35 infectious disease clinical phenotypes implicate broad range of molecular mechanisms

We sought to characterize the genetic determinants of 35 ID traits, including many which have never been investigated using a genome-wide approach. First, we performed GWAS of each of these phenotypes using a cohort of 23,294 and 4,321 BioVU individuals of European and African ancestry, respectively, with extensive EHR information from BioVU^2^. We identified genome-wide significant associations (p < 5×10^−8^) for 13 ID traits (Figure 2A and Supplementary Table 1). The SNP rs17139584 on chromosome 7 was our most significant association (p = 1.21 × 10^−36^) across all traits, with bacterial pneumonia. A LocusZoom plot shows several additional genome-wide significant variants in the locus (Figure 2B), in low linkage disequilibrium (r^2^ < 0.20) with the sentinel variant rs17139584, including variants in the *MET* gene and in *CFTR*. The *MET* gene acts as a receptor to *Listeria monocytogenes* internalin InlB, mediating entry into host cells; interestingly, listeriosis, a bacterial infection caused by this pathogen, can lead to pneumonia^22^. Given the observed associations in the cystic fibrosis gene *CFTR* (∼650 Kb downstream of *MET*), we also asked whether the rs17139584 association was driven by cystic fibrosis. Notably, the SNP remained nominally significant, though its significance was substantially reduced, after adjusting for cystic fibrosis status (p = 0.007; see Methods) or excluding the cystic fibrosis cases (p = 0.02). The LD profile of the genome-wide significant results in this locus (Figure 2B) is consistent with the involvement of multiple independent loci (e.g., *MET* and *CFTR*) underlying bacterial pneumonia risk. The rs17139584 association replicated (p = 5.3×10^−3^) in the UK Biobank^1^. Eighty percent to ninety percent of patients with cystic fibrosis suffer from respiratory failure due to chronic bacterial infection (with *Pseudomonas aeruginosa*)^23^. Thus, future studies on the role of this locus in lung infection associated with cystic fibrosis may provide germline predictors of this complication; alternatively, the locus may confer susceptibility to lung inflammation, regardless of cystic fibrosis status. Collectively, our analysis shows strong support for allelic heterogeneity, with likely multiple independent variants in the locus contributing to interindividual variability in bacterial pneumonia susceptibility.

**Table 1.**
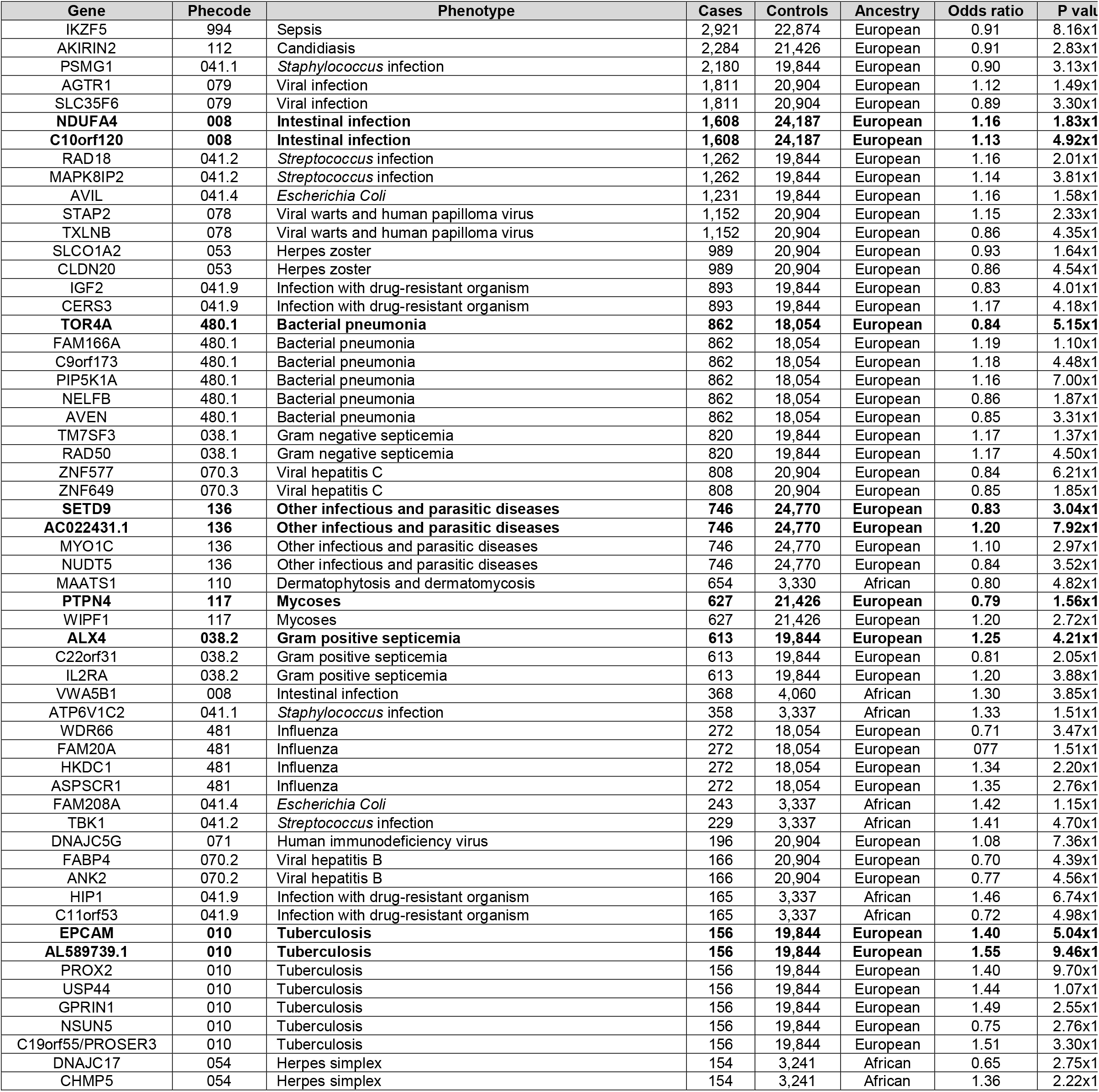
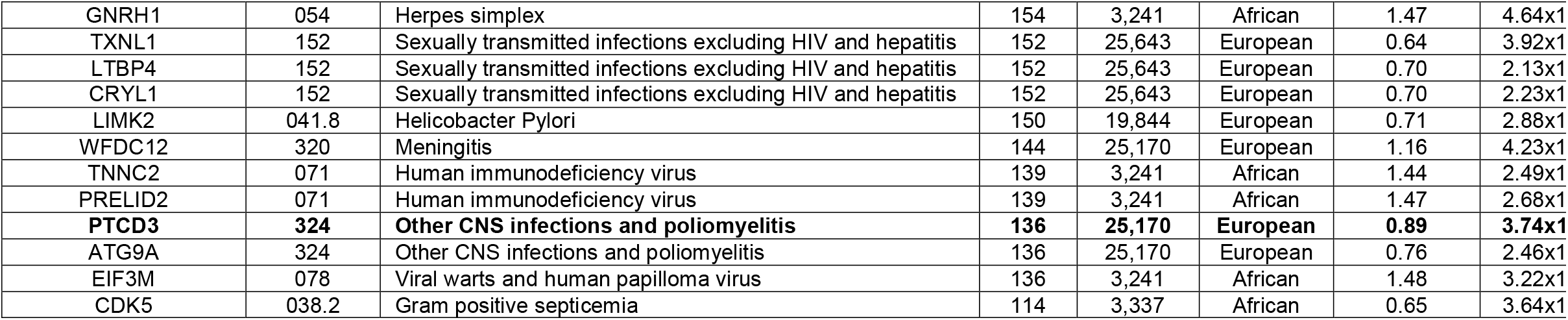
Significant trait-specific gene-level associations with individual infectious disease phenotypes (for which number of cases > 100). Experiment-wide significant findings (adjusted p < 0.05) are noted in **bold**.

**Figure 2.**
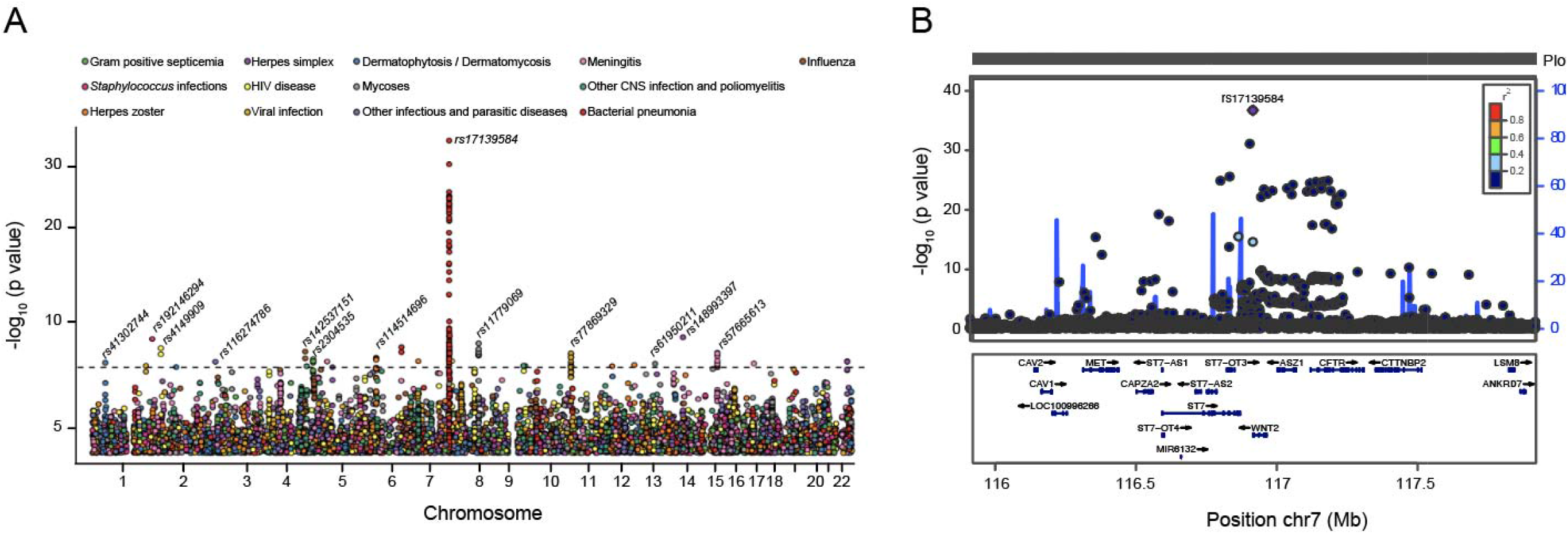
Genome-wide association study (GWAS) of ID traits. (A) Threshold for inclusion of SNP associations was set at 1.0×10^−4^. Genome-wide significance for an ID trait was set at the conventional GWAS threshold p = 5.0×10^−8^, as indicated by the horizontal dotted line. The subset of 13 ID traits (among the full set tested) with variants that meet the traditional genome-wide significance threshold are included. The top variant association for each of the 13 traits is labeled. The most significant variant association is with bacterial pneumonia (p < 1.0×10^−30^). (B) LocusZoom plot at the sentinel variant, rs17139584, associated with bacterial pneumonia. Several variants in low LD (r^2^ < 0.20) with the sentinel variant, including variants in the cystic fibrosis gene *CFTR* and in the *MET* gene >650 Kb upstream, are genome-wide significant for bacterial pneumonia. The sentinel variant remains statistically significant (p = 0.007) after adjusting for a diagnosis of cystic fibrosis.

Additional examples of genome-wide significant associations with other ID traits were identified. For example, rs192146294 on chromosome 1 was significantly associated (p = 1.23×10^−9^) with *Staphylococcus* infection. In addition, 10 variants on chromosome 8 were significantly associated (p < 1.17×10^−8^) with Mycoses infection.

Next, to improve statistical power, we performed multi-tissue PrediXcan^7,14,15^. We constructed an atlas of TWAS associations with these ID traits in separate European and African American ancestry cohorts (Supplementary Data File 1). Notably, 70 genes reached experiment-wide or individual ID-trait significance for 26 of the 35 clinical ID traits (Figure 3A and Table 1). Sepsis, the clinical ID trait with the largest sample size in our data (Figure 3B; Phecode 994; number of cases 2,921; number of controls 22,874), was significantly associated (p = 8.16×10^−7^) with *IKZF5* after Bonferroni correction for the number of genes tested (adjusted p < 0.05). The significant genes (Table 1) were independent of the sentinel variants from the GWAS (Supplementary Table 1), indicating that the gene-based test was identifying additional signals.

**Figure 3.**
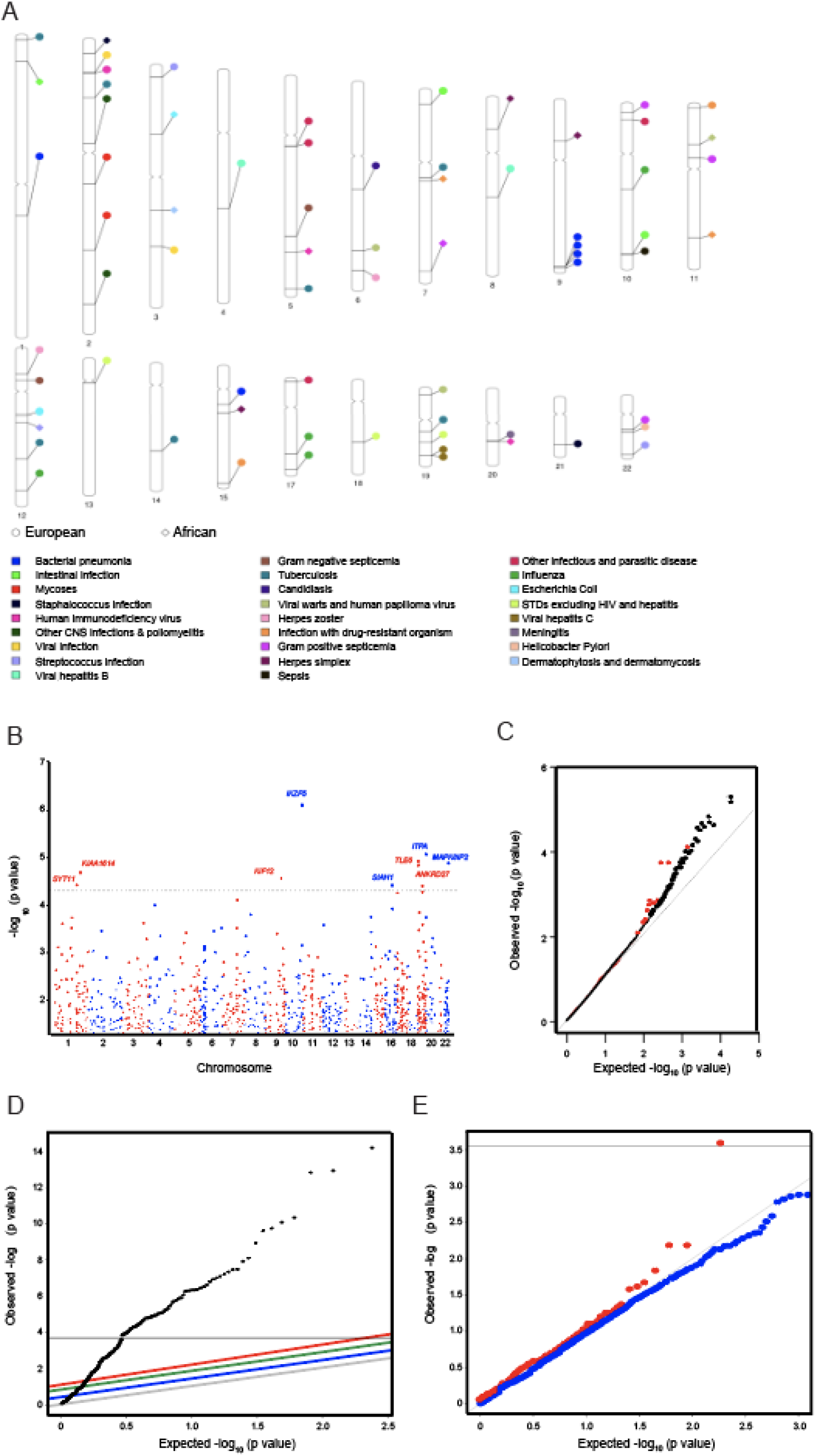
Transcriptome-wide association studies (TWAS) of 35 ID traits reveal novel ID-associated genes. The genetic component of gene expression for autosomal genes was individually tested for association with each of 35 ID traits (see Methods). (A) Experiment-wide or ID-specific significant genes are displayed on the ideogram using their chromosomal locations and color-coded using the associated ID traits. Most associations represent unique genes within the implicated loci, suggesting the genes are not tagging another causal gene. A locus on chromosome 9, by contrast, shows multiple associations with the same ID trait, which may indicate correlation of the expression traits with a single causal gene in the locus. Circle: European ancestry cohort. Diamond: African American ancestry. (B) Manhattan plot shows the PrediXcan associations with sepsis (Phecode 994; number of cases 2,921; number of controls 22,874). Dashed line represents p < 5×10^−5^. The gene *IKZF5* was significant (p = 8.16×10^−7^, adjusted p < 0.05) after Bonferroni correction for the number of genes tested. (C) Q-Q plot of FinnGen replication p-values for genes associated with intestinal infection (p < 0.05) in BioVU (red) compared to the remaining set of genes (black). The ID-associated genes tended to be more significant in the independent dataset than the remaining genes, as evidenced by the leftward shift in the Q-Q plot. (D) Q-Q plot of UK Biobank replication p-values for influenza in lung tissue demonstrating significant deviation from null expectation. The horizontal line corresponds to the Bonferroni threshold (adjusted p < 0.05). (E) Q-Q plot of the full set of FinnGen p-values from the association of gene expression in cerebellum with meningitis (blue) vs. the subset of FinnGen (replication) p-values for the top BioVU meningitis gene-level associations in cerebellum (red). The meningitis associated genes in BioVU dramatically improve signal-to-noise ratio, as shown by the leftward shift in the Q-Q plot. The horizontal line corresponds to the Bonferroni threshold (adjusted p < 0.05) based on the number of top BioVU genes tested.

Our analysis identified previously implicated genes for the specific ID traits but also proposes novel genes and mechanisms. ID-associated genes include *NDUFA4* for intestinal infection, a component of the cytochrome oxidase and regulator of the electron transport chain^24^; *AKIRIN2* for candidiasis, an evolutionarily conserved regulator of inflammatory genes in mammalian innate immune cells^25,26^; *ZNF577* for viral hepatitis C, a gene previously shown to be significantly hypermethylated in hepatitis C related hepatocellular carcinoma^27^; and epithelial cell adhesion molecule (*EPCAM*) for tuberculosis, a known marker for differentiating malignant tuberculous pleurisy^28^, among many others. These examples of ID-associated genes highlight the enormous range of molecular mechanisms that may contribute to susceptibility and complication phenotypes.

### Replication of gene-level associations with infectious diseases in the UK Biobank and FinnGen

To bolster our genetic findings and show that our results were not driven by biobank-specific confounding, we performed extensive replication, using the biobank with the largest sample size (for a given trait) as the discovery dataset (see Methods). Here, replication is defined by concordant direction of effect and statistical significance after Bonferroni adjustment (adjusted p < 0.05) in the test dataset. Replicated genes include *FAM166A* (discovery p = 2.85×10^−11^, replication p = 2.56 ×10^−5^) and *GPATCH11* (discovery p = 2.22×10^−11^, replication p = 4.39 ×10^−3^) for bacterial pneumonia in lung tissue. The complete list of replicated genes by trait and tissue can be found in Supplemental Data File 2.

We investigated the concordance of results across the datasets. For example, we found that the genes associated with intestinal infection (p < 0.05) in BioVU (discovery dataset) – the ID trait with the largest sample size in BioVU and with a matching dataset in the independent FinnGen biobank – showed a significantly greater level of enrichment for gene-level associations with the same trait in FinnGen (test dataset) compared to the remaining set of genes (Figure 3C). In particular, higher significance (i.e., lower p-value) was observed in FinnGen for the intestinal infection associated genes identified in BioVU, which included the top association *NDUFA4* (BioVU p = 1.83×10^−9^, FinnGen p = 0.044). These results suggest there are likely to be additional causal genes among the top associations, which will be identified and replicated when adequate sample sizes for the biobanks are achieved^12,29^.

We identified tissue-level replications for the following ID traits: bacterial pneumonia (lung, Phecode 480.1), influenza (lung, Phecode 481), meningitis (10 brain regions, Phecode 320), and encephalitis (10 brain regions, Phecode 323) (Supplementary Data Files 2-3). As before, to improve statistical power to discover tissue-level associations, we used BioVU as the discovery platform and the UK Biobank as the replication dataset (adjusted p < 0.05, see Methods) (Figure S1A-G and Supplementary Data File 2). This replication analysis identified robust gene-level associations with influenza (Figure 3D, Supplementary Data Files 2-3) and bacterial pneumonia (Figure S1A, Supplementary Data Files 2-3). We provide a longer list of the top genes (p < 0.05 in BioVU) with nominal associations with influenza (Supplementary Data Files 2-3) and viral pneumonia (Supplementary Data File 3) in FinnGen (p < 0.05) as a resource to the community.

We found substantial enrichment for genes with high significance (low p-values) in the respective brain tissues in the UK Biobank (test dataset) for top gene associations with meningitis (p < 0.05) in BioVU (discovery dataset) in hippocampus, cerebellar hemisphere, and hypothalamus (Figure S1B-D, Supplementary Data Files 2-3). Similar enrichment patterns were observed for the remaining brain tissues (Supplementary Data Files 2-3). Likewise, significant enrichment results (Figure S1E-G, Supplementary Data Files 2-3) in brain tissues (with BioVU as discovery dataset and UK Biobank as test dataset) were observed for encephalitis. Using FinnGen (test dataset), we continued to observe enrichment (though not as pronounced) for genes with high significance for some ID traits. For example, the top cerebellar gene associations with meningitis (p < 0.05) in BioVU improved the signal-to-noise ratio for the cerebellar associations in FinnGen (Figure 3E).

### Tissue expression profile of infectious disease associated genes suggests tissue-dependent mechanisms

The ID-associated genes tend to be less tissue-specific (i.e., more ubiquitously expressed) than the remaining genes (Figure S2A, Mann Whitney U test on the τ statistic, p = 7.5×10^−4^), possibly reflecting the multi-tissue PrediXcan approach we implemented, which prioritizes genes with multi-tissue support to improve statistical power. We hypothesized that tissue expression profiling of ID-associated genes can provide additional insights into disease etiologies and mechanisms. For example, the intestinal infection associated gene *NDUFA4* is expressed in a broad set of tissues, including the alimentary canal, but displays relatively low expression in whole blood (Figure S2B). In addition, *TOR4A*, the most significant association with bacterial pneumonia (Table 1), is most abundantly expressed in lung, consistent with the tissue of pathology, but also in spleen (Figure S2C), whose rupture is a lethal complication of the disease^30,31^. These examples illustrate the diversity of tissue-dependent mechanisms that may contribute in complex and dynamic ways to interindividual variability in ID susceptibility and progression. We therefore provide a resource of gene-level associations with the ID traits in a broad collection of tissues to facilitate molecular or clinical follow-up studies.

### Genetic overlap reveals host gene expression programs and common pathways as targets for pathogenicity

We hypothesized that ID-associated genes converge on shared functions and pathways, which may reflect common targeted host transcriptional programs. Among the 70 gene-level associations with the 35 clinical ID traits, 40 proteins are post-translationally modified by phosphorylation (Supplementary Table 2), a significant enrichment (Benjamini-Hochberg adjusted p < 0.10 on DAVID annotations^32^) relative to the rest of the genome, indicating that phosphoproteomic profiling can shed substantial light on activated host factors and perturbed signal transduction pathways during infection^33,34^. In addition, 16 proteins are acetylated, consistent with emerging evidence supporting this mechanism in the host antiviral response^35^ (Supplementary Table 3). These data highlight specific molecular mechanisms across ID traits with critical regulatory roles (e.g., protein modifications) in host response among the ID-associated genes.

We tested the hypothesis that distinct infectious agents exploit common pathways to find a compatible intracellular niche in the host, potentially implicating shared genetic risk factors. Notably, 64 of the 70 ID-associated genes (Table 1) were nominally associated (p < 0.05) with multiple ID traits (Supplementary Table 4). These genes warrant further functional study as broadly exploited mechanisms targeted by pathogens or as broadly critical to pathogen-elicited immune response. Gene Set Enrichment Analysis (GSEA) of these genes implicated a number of significant (FDR < 0.05) gene sets (Figure 4A), including those involved in actin-based processes and cytoskeletal protein binding, processes previously demonstrated to mediate host response to pathogen infection^36^. Since diverse bacterial and viral pathogens target host regulators that control the cytoskeleton (which plays a key role in the biology of infection) or modify actin in order to increase virulence, intracellular motility, or intercellular spread^37-39^, these results reassuringly lend support to the involvement of the genes in infectious pathogenesis.

**Figure 4.**
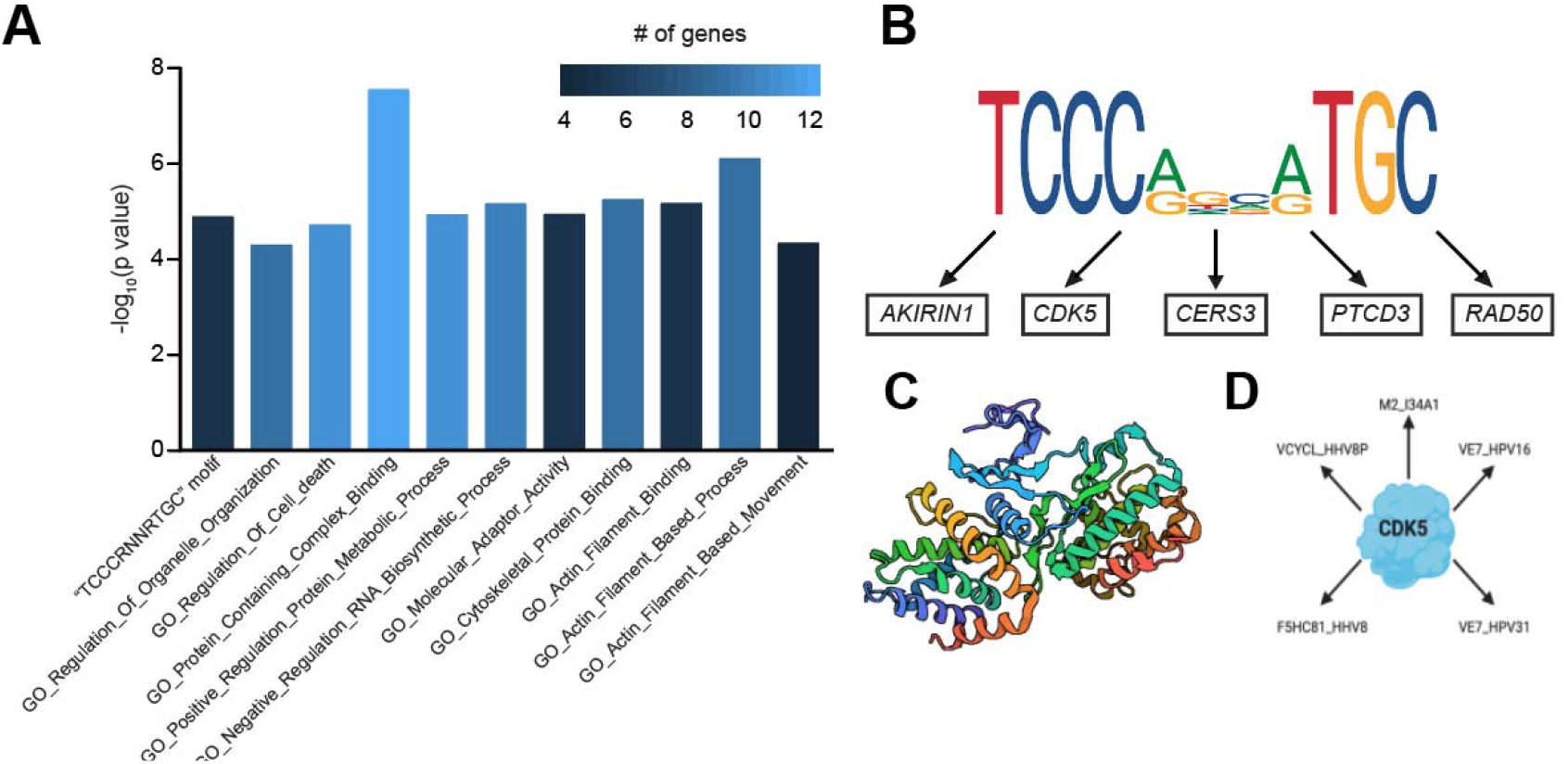
Enriched pathways across multiple ID traits and pathogen evolutionary strategies to promote infection. (A) Gene set enrichment analysis of ID associated genes having also nominal associations with additional ID traits. All gene sets satisfied false discovery rate < 0.05 for pathway enrichment and included known biological processes (e.g. protein complex formation, cytoskeletal protein binding, cell death, actin motility, etc.) relevant to the biology of infection. (B) Highly conserved motif “TCCCRNNRTGC”, within 4 kb of the transcription start site (TSS) of ID-associated genes, is enriched among the multi-ID associated genes and does not match any known transcription factor binding site. Genes with this motif near the TSS include *AKIRIN2, CDK5, RAD50, PTCD3*, and *CERS3*. This suggests a strategy that the pathogens may broadly exploit to hijack the host transcriptional machinery. (C) CDK5 is an example of a multi-ID associated gene, significantly associated with Gram-positive septicemia and nominally associated with other IDs, including herpes simplex virus. CDK5 is activated by its regulatory subunit p35/p25. The CDK5-p25 complex regulates inflammation (whose large-scale disruption is characteristic of septicemia) and induces cytoskeletal disruption in neurons (where the herpes virus promotes lifelong latent infection). Structure of the CDK5-p25 complex (PDB: 1H4L, ^44^) is shown here. The A and B chains are required for cytoskeletal protein binding (CDK5), whereas the D and E chains (p25) are involved in actin regulation and kinase function, all functions implicated in our pathway analysis. (D) Multi-ID associated genes identified by our study have also been observed in host-pathogen protein complexes (by coimmunoprecipitation, affinity chromatography, and two-hybrid approaches, among others) for the specific pathogens responsible for the ID traits. Interactions of pathogen proteins with CDK5 are shown here. M2_134A1 (UniProt: PO6821) is the matrix protein 2 component of the proton-selective ion channel required for influenza A viral genome release during cellular entry and is targeted by the anti-viral drug amantadine^48^. VE7_HPV16 (UniProt: PO3129) is a component of human papillomavirus (HPV) required for cellular transformation and trans-activation through disassembly of E2F1 transcription factor from RB1 leading to impaired production of type I interferons^49-51^. VE7_HPV31 (UnitProt: P17387) engages histone deacetylases 1 and 2 to promote HPV31 genome maintenance^52^. VCYCL_HHV8P (UniProt: Q77Q36) is a cyclin homolog within the human herpesvirus 8 genome that has been shown to control cell cycle through CDK6 and induce apoptosis through Bcl2^53-55^. F5HC81_HHV8 (UniProt: F5HC81) is not well-characterized, but predicted to act as a viral cyclin homolog. This suggests a second strategy that the pathogens exploit, i.e., alteration of the host proteome, to promote infection.

Notably, we identified an enrichment (FDR = 9.68×10^−3^) for a highly conserved motif (“TCCCRNNRTGC”), within 4 kb of transcription start site (TSS) of multi-ID associated genes (Figure 4A-B), that does not match any known transcription factor binding site^40^ and may be pivotal for host-pathogen interaction for the diversity of infectious agents included in our study. In addition, we found that several of the multi-ID associated genes (with the sequence motif near the TSS) have been observed in host-pathogen protein complexes (by both coimmunoprecipitation and affinity chromatography approaches) for the specific pathogens responsible for the ID traits^41^. See Supplementary Data File 4 for complete list of host-pathogen interactions for these genes/proteins. One example is *CDK5*, a gene significantly associated with Gram-positive septicemia (Table 1) and nominally associated with multiple ID traits, including herpes simplex. CDK5 is activated by p35, whose cleaved form p25 results in subcellular relocalization of CDK5. The CDK5-p25 complex regulates inflammation^42^ (whose large-scale disruption is characteristic of septicemia) and induces cytoskeletal disruption in neurons^43^ (where the herpes virus is responsible for lifelong latent infection). The A and B chains of the CDK5-p25 complex (Figure 4C for structure diagram^44^) are required for cytoskeletal protein binding (CDK5), whereas the D and E chains (p25) are involved in actin regulation and kinase function, all molecular processes implicated in our pathway analysis. Intriguingly, blocking CDK5 can have a substantial impact on the outcome of inflammatory diseases including sepsis^45^, enhancing the anti-inflammatory potential of immunosuppressive treatments, and has been shown to attenuate herpes virus replication^46^, suggesting that modulation of this complex is important for viral pathogenesis.

CDK5 is also altered by several other viruses, identified using unbiased mass spectrometry analysis^47^ (Figure 4D), indicating a broadly exploited mechanism (across pathogens) that is consistent with the gene’s multi-ID genetic associations in our TWAS data (Figure 4D). The CDK5-interaction proteins include: 1) M2_134A1 (matrix protein 2, influenza A virus), a component of the proton-selective ion channel required for viral genome release during cellular entry and is targeted by the anti-viral drug amantadine^48^; 2) VE7_HPV16, a component of human papillomavirus (HPV) required for cellular transformation and trans-activation through disassembly of E2F1 transcription factor from RB1 leading to impaired production of type I interferons^49-51^; 3) VE7_HPV31, which has been shown to engage histone deacetylases 1 and 2 to promote HPV31 genome maintenance^52^; 4) VCYCL_HHV8P (cyclin homolog within the human herpesvirus 8 genome), which has been shown to control cell cycle through CDK6 and induce apoptosis through Bcl2^53-55^; and 5) F5HC81_HHV8, predicted to act as a viral cyclin homolog. Overall, these data underscore the evolutionary strategies that pathogens have evolved to promote infection, including the hijacking of the host transcriptional machinery and the biochemical alterations of the host proteome.

### Serology and culture data reveal insights into clinical infection and pathogen colonization

We exploited extensive clinical microbiological laboratory analysis of blood (Figure 5A), bronchoalveolar lavage, sputum, sinus/nasopharyngeal, and tracheal cultures for bacterial and fungal pathogen genus identification (Figure S3A-D), as well as respiratory viral genus identification (Figure S3E) (see Methods) to evaluate phenotype resolution and algorithm. For example, we found that *Staphylococcus* infection (Phecode = 041.1) performed well in classifying *Staphylococcus aureus* infection based on blood culture data. The area under the Receiver Operating Characteristic (ROC) curve was 0.938 (Figure 5B) with standard error of 0.008 generated from bootstrapping (see Methods). The area under the curve (AUC) quantifies the probability that the Phecode classifier ranks a randomly chosen positive instance of *Staphylococcus aureus* infection in blood higher than a randomly chosen negative one. In comparison, the first principal component (PC) in our European ancestry samples showed AUC of 0.514 (Figure 5B) while sex and age performed even more poorly (AUC 0.50). We then tested a logistic model with the Phecode classifier, age, sex, and the first 5 PCs in the model. The Phecode classifier was significantly associated (p < 2.2×10^−16^) after conditioning on the remaining covariates. The fitted value from the joint model consisting of the remaining covariates showed AUC of 0.568 (Figure 5B). Collectively, culture data for improved resolution of clinical infection and pathogen colonization provide validation of our approach.

**Figure 5.**
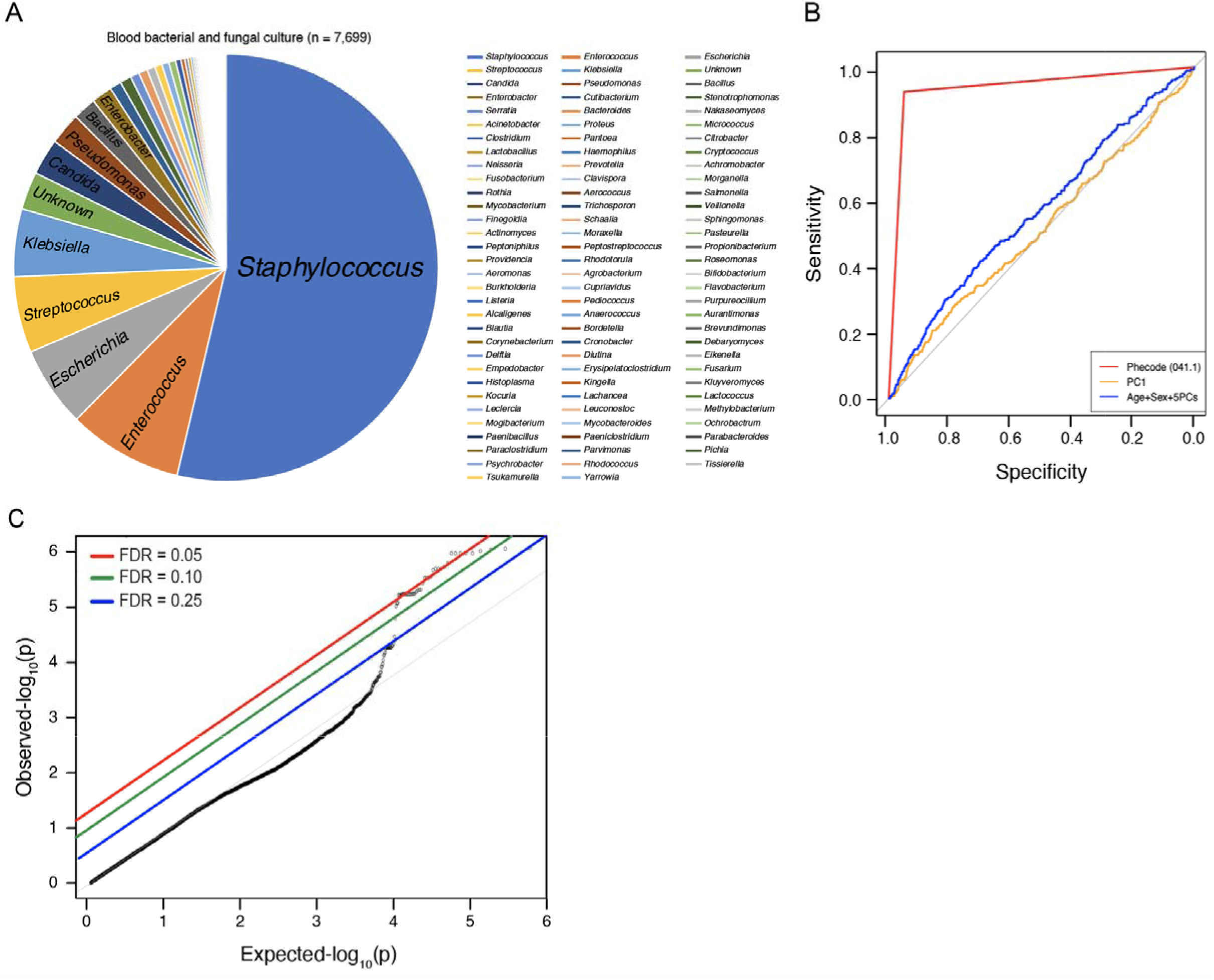
Pathogen genus identification from gut microbiome and clinical blood cultures linked to whole-genome information reveals insights into host colonization and infection. (A) Bacterial and fungal pathogens identified from blood (n = 7,699 positive cultures across 94 genera) from 2,417 individuals. (B) Area under the receiver operating characteristic curve (AUC) showing that the clinical trait *Staphylococcus* infection (Phecode = 041.1) performs well in classifying *Staphylococcus aureus* infection based on blood culture data from (A), with AUC of 0.938 with standard error of 0.008. The first PC in the European ancestry samples and a model with age, sex, and the first 5 PCs, both with substantially lower performance (AUC of 0.514 and 0.568, respectively), are also shown. (C) Q-Q plot of top SNPs (p < 0.0001) associated with *Desulfovibrionaceae* colonization in the human gut microbiome showing the p-value distribution from the association with intestinal infection (Phecode 008) in BioVU. Note the enrichment as shown by the leftward shift relative to the diagonal line, with several genes satisfying the FDR<0.05 (red line) threshold.

To expand these findings and further dissect the complex pathogen-colonization patterns in humans, we utilized host genome-wide associations with human gut microbiome variation for 155 pathogens^17^ identified through 16S rRNA sequencing. For example, among the top SNP associations with *Desulfovibrionaceae* (p<0.0001), a sulfate-reduced bacterium associated with intraabdominal infections and inflammatory bowel disease^56,57^, we observed an enrichment for SNPs associated with intestinal infection in BioVU (Figure 5C). These data provide a reference resource to explore how genetically-determined microbiome variation influences ID trait susceptibility.

### Phenome scan of clinical ID-associated genes identifies adverse outcomes and complications

Electronic Health Records (EHR) linked to genetic data may reveal insights into associated clinical sequalae^58-60^. To assess the phenomic impact of ID-associated genes (Table 1), we performed a phenome-scale scan across 197 hematologic, respiratory, cardiovascular, and neurologic traits available in BioVU (Figure 6A and Supplementary Data File 5). Correcting for total number of genes and phenotypes tested, we identified four gene-phenotype pairs reaching experiment-wide significance (adjusted p < 0.05): 1) *WFDC12*, our most significant (p = 4.23×10^−6^) association with meningitis and a known anti-bacterial gene^61^, is also associated with cerebral edema and compression of brain (p = 1.35×10^−6^), a feared clinical complication of meningitis^62^; 2) *TM7SF3*, the most significant gene with Gram-negative sepsis (p = 1.37×10^−6^), is also associated with acidosis (p = 1.95×10^−6^), a known metabolic derangement associated with severe sepsis^63^, and a gene known to play a role in cell stress and the unfolded protein response^64^; 3) *TXLNB*, the most significant gene associated with viral warts and human papillomavirus infection (p = 4.35×10^−6^), is also associated with abnormal involuntary movements, p = 1.39×10^−6^; and 4) *RAD18*, the most significant gene associated with Streptococcus infection (p = 2.01×10^−6^), is also associated with anemia in neoplastic disease (p = 3.10×10^−6^). Thus, coupling genetic analysis to EHR data with their characteristic breadth of clinical traits offers the possibility of determining the phenotypic consequences of ID-associated genes, including known (in the case of *WFDC12* and *TM7SF3*) potentially adverse health outcomes and complications.

**Figure 6.**
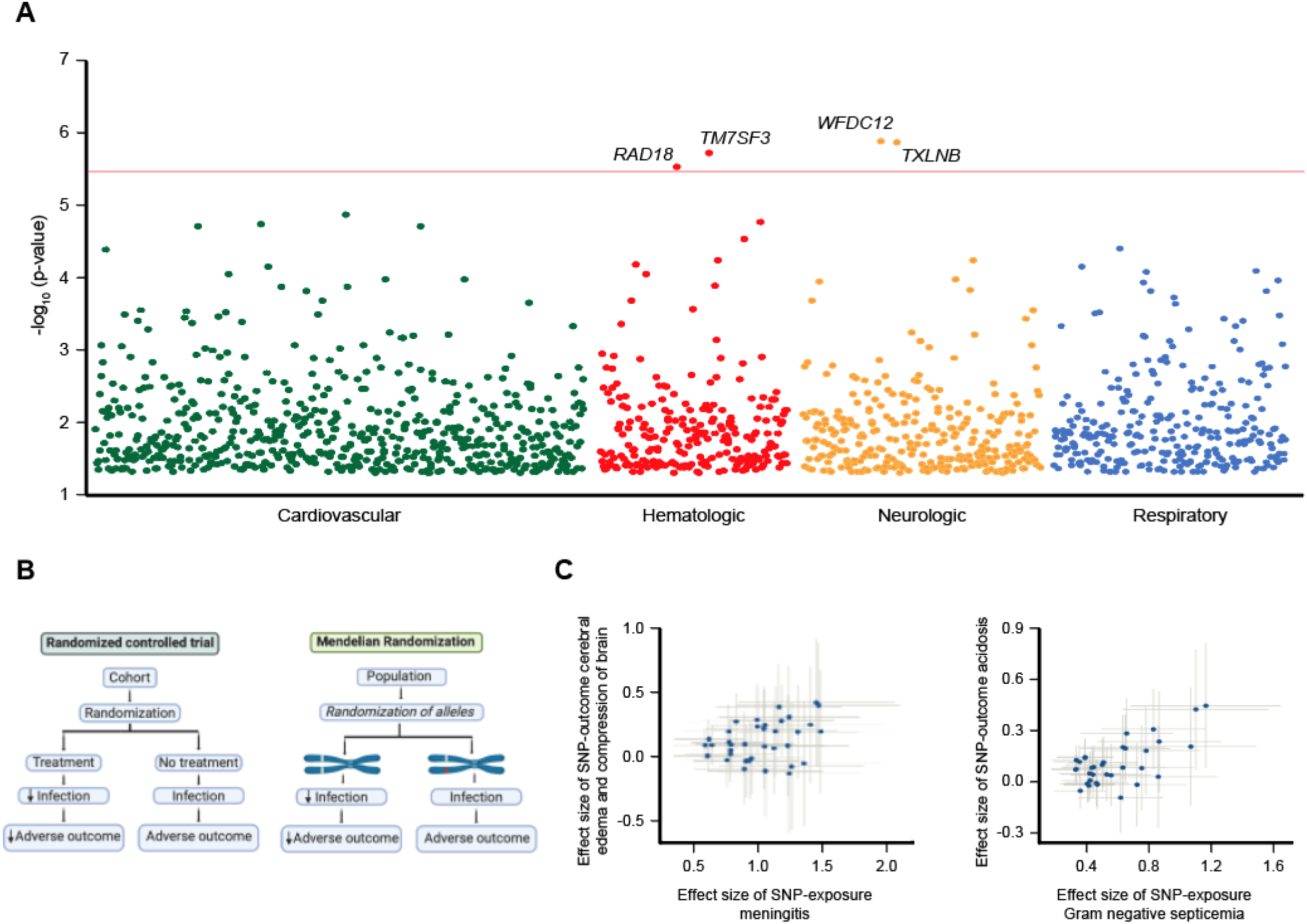
Phenome-scale scan of 70 ID-associated genes across 197 cardiovascular, hematologic, neurologic, and respiratory phenotypes (cases > 200) in BioVU (phewascatalog.org) identifies genes association with both disease risk and corresponding known complications of the infection. (A) Each dot represents the association of an ID-associated gene with one of the 197 (hematologic, respiratory, cardiovascular, and neurologic) phenotypes. Horizontal red line indicates threshold for statistical significance correcting for number of phenotypes and ID-associated genes tested. We identify four gene-phenotype pairs reaching experiment-wide significance (adjusted p < 0.05): 1) *WFDC12*, our most significant (p = 4.23×10^−6^) association with meningitis, is also associated with cerebral edema and compression of brain (p = 1.35×10^−6^), a feared clinical complication of meningitis^62^; 2) *TM7SF3*, the most significant gene with Gram-negative sepsis (p = 1.37×10^−6^), is also associated with acidosis (p = 1.95×10^−6^), a known metabolic derangement associated with severe sepsis^63^; 3) *TXLNB*, the most significant gene associated with viral warts and human papillomavirus infection (p = 4.35×10^−6^), is also associated with abnormal involuntary movements, p = 1.39×10^−6^; and 4) *RAD18*, the most significant gene associated with Streptococcus infection (p = 2.01×10^−6^), is also associated with anemia in neoplastic disease (p = 3.10×10^−6^). (B) Mendelian randomization framework. P-value threshold used to define an instrumental variable was set at p < 1.0×10^−5^ and variants in linkage equilibrium (r^2^ = 0.01) were used. (C) Mendelian Randomization provides strong support for causal exposure-outcome relationships for 1) meningitis and compression of brain (left, median-weighted estimator p = 2.7×10^−3^); and 2) gram-negative septicemia and acidosis (right, median-weighted estimator p = 2.0×10^−7^). Grey lines indicate 95% confidence interval.

### Mendelian Randomization provides causal support for the effect of infectious disease trait on identified adverse phenotypic outcomes/complications

Since our gene-level associations with clinical ID diagnoses implicated known adverse complications, we sought to explicitly evaluate the causal relation between the ID traits and the adverse outcomes/complications. We utilized the Mendelian Randomization paradigm^18^ (Figure 6B), which exploits genetic instruments to make causal inferences in observational data, in effect, performing randomized controlled trials to evaluate the causal effect of “exposure” (i.e., ID trait) on “outcome” (e.g., the complication). Specifically, we conducted multiple-instrumental-variable causal inference using GWAS ^21^ and PrediXcan summary results. First, we used independent SNPs (r^2^ = 0.01) with association p < 1.0×10^−5^ as genetic instruments. To control for horizontal pleiotropy and account for the presence of invalid genetic instruments, we utilized MR-Egger regression and weighted-median Mendelian Randomization (see Methods)^65,66^.

We found causal support for the effect of 1) Gram-negative sepsis on acidosis (Figure 6C, weighted-median estimator p = 2.0×10^−7^); and 2) meningitis on cerebral edema and compression of brain (Figure 5C, weighted-median estimator p = 2.7×10^−3^). Our resource establishes a platform to elucidate the genetic component of an ID trait and its impact on the human disease phenome, enabling causal inference on the effect of an ID trait on potential complications.

### TWAS of 79 pathogen-exposure induced cellular traits highlights cellular mechanisms and enables validation of ID gene-level associations

Elucidating how the genes influence infection-related cellular trait variation may provide a mechanistic link to ID susceptibility. We thus performed TWAS of 79 pathogen-induced cellular traits – including infectivity and replication, cytokine levels, and host cell death, among others^19^ (Supplementary Data File 6). We identified 38 gene-level associations reaching trait-level significance (p < 2.87×10^−6^, correcting for number of statistical tests; Figure 7A). In addition, we replicated SNP associations with the cellular traits using the genetic associations with the ID traits (Supplementary Data File 7) that map to the specific cellular phenotypes (Supplementary Data File 8).

**Figure 7.**
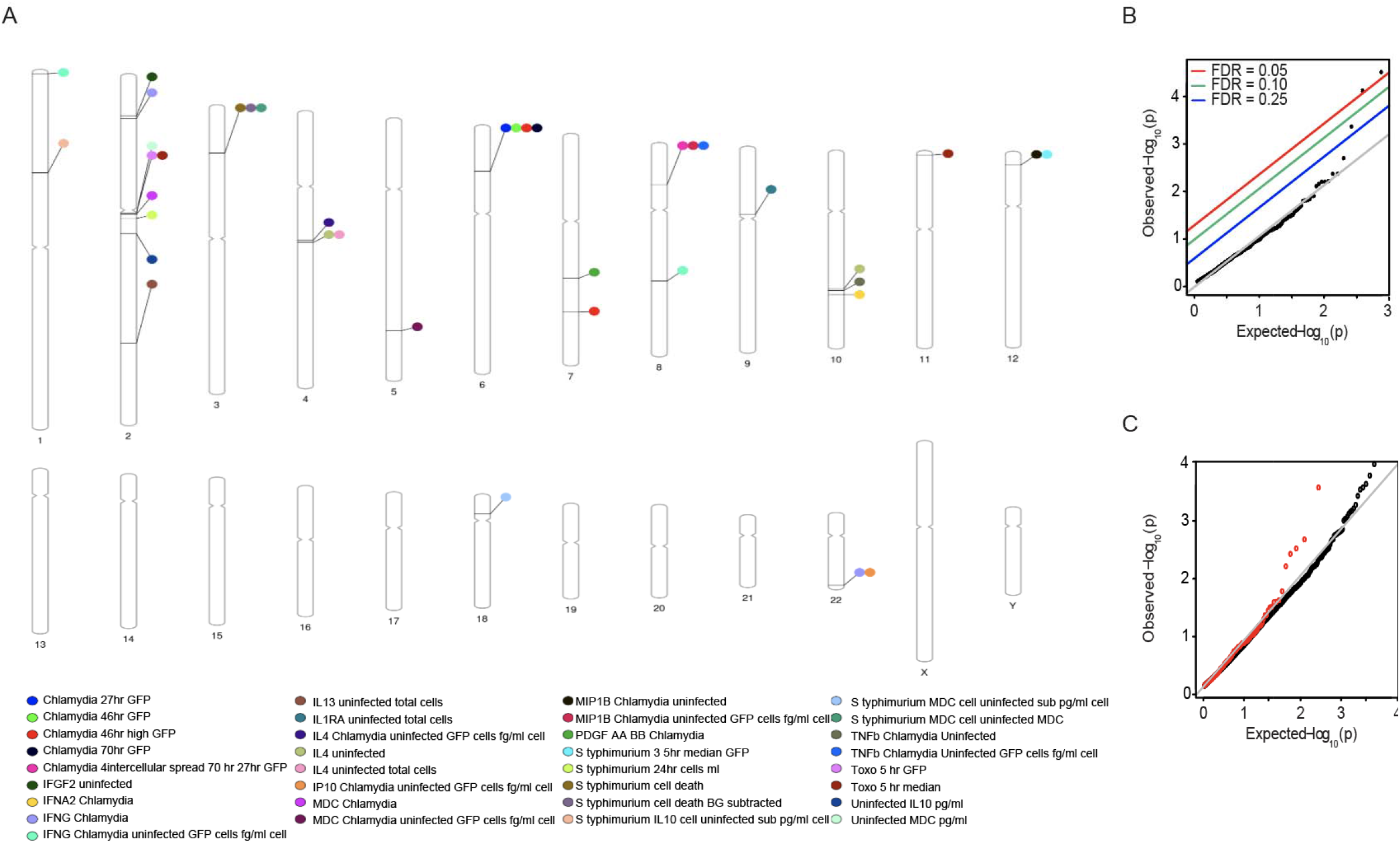
TWAS of 79 pathogen-exposure induced cellular traits improves identification of pathogen-induced cellular mechanisms. (A) Genes reaching significance in Hi-HOST after correction for the total number of genes and cellular phenotypes tested. (B) Integration of EHR data into Hi-HOST facilitates replication of gene-level associations with a clinical ID trait. Genes nominally associated (p < 0.05) with Gram-positive septicemia (Phecode 038.2) in BioVU show significant enrichment for *Staphylococcus* toxin exposure, a Hi-HOST phenotype. The Q-Q plot shows the distribution of TWAS p-values in the Hi-HOST data for the top genes in the BioVU data. False discovery rate (FDR) thresholds at 0.25 (blue), 0.10 (green), and 0.05 (red) are shown. (C) Integration of EHR data into Hi-HOST also improves the signal-to-noise ratio in Hi-HOST. For example, the top 300 genes nominally associated with *Staphylococcus* infection (Phecode 041.1) in BioVU (p < 0.016, red) depart from null expectation for their TWAS associations with *Staphylococcus* toxin exposure in Hi-HOST compared to the full set of genes (black).

Integration of EHR data into Hi-HOST^19^ may provide additional functional support for the gene-level associations with a clinical ID trait. For example, we observed a marked enrichment for genes associated with direct *Staphylococcus* toxin exposure cellular response in Hi-HOST among the human Gram-positive septicemia associated genes from BioVU (see Supplementary Data File 6 for genes with FDR < 0.05) (Figure 7B). In addition, integration of EHR data into Hi-HOST may improve the signal-to-noise ratio in Hi-HOST TWAS data. Indeed, the top 300 genes nominally associated with Staphylococcus infection (Phecode 041.1) in BioVU departed from null expectation for their associations with Staphylococcus toxin exposure in Hi-HOST compared to the full set of genes, which did not (Figure 7C), as perhaps expected due to the modest sample size. Collectively, these results demonstrate that integrating the EHR-derived TWAS results into TWAS of the cellular traits can greatly improve identification of potentially relevant pathogenic mechanisms.

### Phenome scan of TWAS findings from Hi-HOST

To identify potential adverse effects of direct pathogen exposure, we performed a phenome-scan across the 197 cardiovascular, hematologic, neurologic, and respiratory traits as described above. Our top gene-phenotype pairs include: 1) *FAM171B*, our most significant association with interleukin 13 (IL-13) levels is also associated with alveolar and parietoalveolar pneumonopathy (p = 4.04×10^−5^), a phenotype known to be modulated by IL-13 dependent signaling^67^; 2) *OSBPL10*, the most significant gene associated with cell death caused by *Salmonella enterica serovar Typhimurium*, is also associated with intracerebral hemorrhage (p = 4.99×10^−5^), a known complication of *S. Typhimurium* endocarditis^68^. These data highlight the utility of joint genetic analysis of pathogen-exposure-induced phenotypes and clinical ID traits to gain insights into the molecular and cellular basis of complications and adverse outcomes. However, more definitive conclusions will require larger sample sizes and functional studies.

## DISCUSSION

ID susceptibility is a complex interplay between host genetic variation and pathogen-exposure induced mechanisms. While GWAS has begun to identify population-dependent loci conferring ID risk^69^, the underlying function of identified variants, predominantly in non-coding regulatory regions, remains poorly understood. Molecular characterization of infectious processes has been, in general, agnostic to the genetic architecture of clinical infection. Although pathogen exposure is requisite to display clinical ID traits, characterizing the role of host genetic variation remains challenging.

Our study provides a reference atlas of genetic variants and genetically-determined expression traits associated with a diverse set of clinical ID traits. We identified 70 gene-level associations in BioVU, with replication for a subset of ID traits in the UK Biobank and FinnGen. To provide additional support to our findings, we leveraged a rich resource of genetic information linked to serologic tests and pathogen cultures from five clinical sample sites and exploited a large catalog of genome-wide associations of microbiome variation generated from 16S rRNA based taxonomic classification. A phenome scan across 197 hematologic, respiratory, cardiovascular, and neurologic traits proposes a molecular basis for the link between certain ID traits and outcomes. Using Mendelian Randomization, we determined the ID traits which, as exposure, show significant causal effect on outcomes. Finally, we developed a TWAS catalog of 79 pathogen-exposure induced cellular traits (Hi-HOST) in a broad collection of tissues, which provides a platform to interrogate mediating cellular and molecular mechanisms.

Genetic predisposition to ID onset and progression is likely to be complex^70^. Monogenic mechanisms conferring ID risk have been proposed, but these mechanisms are unlikely to explain the broad contribution of host genetic influence on ID risk^71^. Thus, a function-centric methodology is necessary to disentangle potentially causal pathways. Our approach builds on PrediXcan, which estimates the genetically-determined component of gene expression^14^. The genetic component of gene expression can then be tested for association with the trait, enabling insights into potential pathogenic mechanisms^4^ and novel therapeutic strategies^72^. Leveraging the shared regulatory architecture of gene expression^7^ can help prioritize gene mechanisms for downstream functional studies to provide an avenue to identify causal cell types and genes. Our results highlight a multi-tissue approach to infer causal genes and pathways relevant to ID biology.

Our study identified genes with diverse functions, including roles in mitochondrial bioenergetics^24,73^, regulation of cell death^74^, and of course links to host immune response^75-84^. These diverse functions may contribute to pleiotropic effects on clinical outcomes and complications. In addition, we identified genes implicated in Mendelian diseases, for which susceptibility to infection is a predominant feature, including *WIPF1* (OMIM #614493; recurrent infections and reduced natural killer cell activity^85^), *IL2RA* (OMIM #606367; recurrent bacterial infections, recurrent viral infections, and recurrent fungal infections^79^), and *TBK1* (OMIM #617900; herpes simplex encephalitis (HSE), acute infection, and episodic HSE^86^). These examples show that the identified genes may also confer predisposition, with near-complete penetrance, to an infectious disease related trait displaying true Mendelian segregation.

Enrichment analysis of 64 of the 70 ID-associated genes with nominal support for associations with other clinical ID traits identified modulation of the actin cytoskeleton as a potential shared mechanism of host susceptibility to infection (Figure 4). While manipulation of the actin cytoskeleton by pathogens is hardly a new concept, our study identified specific host genetic variation in actin regulatory genes that is potentially causative of clinical ID manifestations. In addition to pathogen interaction with the cytoskeletal transport machinery, efficient exploitation of host gene expression program is crucial for successful invasion and colonization, and here we mapped several pathogenicity-relevant targets. Notably, we observed a significant enrichment for a highly conserved sequence motif, within 4 kb of a multi-ID-associated gene’s TSS, that is not a known transcription factor binding site. The motif’s presence near multi-ID associated genes suggests a broad regulatory role in host-pathogen interaction, involving the diversity of pathogens examined here, towards successful reprogramming of host gene expression. Furthermore, we identified a significant enrichment for phosphorylated host proteins, suggesting the value of global phosphoproteomic profiling, which has recently been used to prioritize pharmacological targets for the novel SARS-CoV-2 virus^87^. These data highlight several potential avenues by which host susceptibility can be breached by a pathogen’s requirement to maintain a niche through manipulation of host cellular machinery.

To obtain additional support for our gene-level associations, we leveraged two genomic resources with rich phenotypic information (UK Biobank^1^ and FinnGen^16^). These data will prove increasingly useful to characterizing the genetic basis of the ID-associated adverse outcomes and complications. Despite the caveats for the use of EHR in genetic analyses of ID traits^12,29^, the growing availability of such independent datasets will facilitate identification of robust genetic associations. Perhaps more importantly, the breadth of clinical phenotypes in these EHR datasets should enable identification of associated adverse outcomes and complications for the ID-associated genes.

The primary challenges in conducting GWAS of ID traits include phenotype definition and case-control misclassification. Obstacles to accurate phenotype definition include the requirement of specialized laboratory testing to identify specific pathogens and administration of prophylactic therapeutics complicating identification of potentially causative pathogens. Seropositivity may result from the complex genetic properties of the pathogen and the particular mechanisms governing host-pathogen interaction. However, seropositivity may not indicate clinical manifestations of the disease. On the other hand, seronegativity may imply lack of exposure to the pathogen, the absence of infection even in the presence of exposure, or host resistance to infection. Anchoring the analysis to host genetic information (as in our use of genetically-determined expression) and replication of discovered associations may address some aspects of this challenge. Here we exploit an extensive resource of culture data (for identification of pathogens from clinical specimens) linked to whole-genome genetic information to provide additional support to our gene-level associations. One of the ubiquitous problems in diagnosis is that culture recovery is often for multiple organisms, or a contaminant not relevant to the actual pathogen. Similarly, molecular diagnostics of pathogen identification is often a curation of multiple statistically relevant putative pathogens. The mapping of pathogen genome identification to transcriptional response (molecular seropositivity) is a valuable validation of a finding that a given pathogen is associated with a particular infectious syndrome, and our approach to identification of genetically-determined expression changes may facilitate this mapping. Future studies may also implement more complex GWAS models, including incorporating the pathogen genome.

The catalog of TWAS associations with microbiome composition may facilitate insights into molecular mechanisms of infectious disease risk and complications, inform studies of host-pathogen interactions, and improve anti-microbial pharmacologic strategies. Improved characterization of pathogen colonization and taxonomic classification at species and strain level through 16S rRNA sequencing-based approaches may lead to greater resolution of causative infectious processes. Disruption of pathogen equilibrium in the microbiome by environmental or genetic variation may determine susceptibility to human disease^88,89^. However, critical challenges to understanding the patterns of host colonization include identification of rare pathogen populations as well as environmental pressures (i.e. medication use, dietary alterations, etc.) acutely altering the microbiome landscape^90^. Thus, linking microbiome traits to host genetic variation promises to improve resolution of causative mechanisms for ID traits and potentially adverse outcomes.

Mendelian Randomization provides a framework to perform causal interference on the effect of the exposure on the outcome^18,21^ using genetic instruments. We leveraged a summary statistics based approach to test the causal effect of an ID trait on potential adverse outcomes. Mendelian Randomization requires three assumptions: 1) the genetic instrument is associated with exposure (i.e., ID trait); 2) the genetic instrument is associated with the outcome (i.e., adverse outcome or complication) only through the exposure of interest; and 3) the genetic instrument is affecting the outcome independent of other factors (i.e., confounders). Violations of these assumptions can have critical implications for the interpretation of the results. Thus, several approaches have been developed that are robust to these violations. In the case of ID traits, a methodology that distinguishes causality from comorbidity is critical. While many phenotypes are highly comorbid and suspected to have a causal relationship (e.g., smoking and depression/anxiety), Mendelian Randomization does not necessarily support the causal hypothesis^91^. Furthermore, since RCTs cannot be ethically conducted for ID traits and adverse outcomes, the methodology offers an approach for elucidating the role of an infection phenotype or pathogen exposure in disease causation using an observational study design. Here, we found strong causal support for the effect of certain clinical ID traits on potential adverse complications identified through a phenome scan of the ID-associated genes: 1) meningitis -cerebral edema and compression of brain; and 2) Gram-negative sepsis -acidosis.

To enable investigations into mediating cellular and molecular traits for the ID-associated genes, we provide a functional genomics resource built on a high-throughput *in vitro* pathogen infection screen (Hi-HOST)^19^. Integration of EHR data into Hi-HOST facilitates replication of gene-level associations with clinical ID traits and greatly improves the signal-to-noise ratio. This discovery and replication platform, encompassing human phenomics and cellular microbiology, provides a high-throughput approach to linking host cellular processes to clinical ID traits and adverse outcomes.

Although additional mechanistic studies are warranted, our study provides a foundation for anchoring targeted molecular studies in human genetic variation. Elucidation of host mechanisms exploited by pathogens requires multi-disciplinary approaches. Here, we show the broader role of host genetic variation, implicating diverse disease mechanisms. Our study generates a rich resource and exploits a genetics-anchored methodology to facilitate investigations of ID-associated clinical outcomes and complications. Causal inference on the clinical ID traits and potential complications promises to expand our understanding of the molecular basis for the link and, crucially, enable prediction and prevention of serious adverse events.

## METHODS

### BioVU

BioVU, one of the largest DNA biobanks tied to an EHR database, is a subset of the synthetic derivative (SD), a deidentified electronic health record, consisting of individuals with whole-genome genetic information. Detailed information on the construction, utilization, ethics, and policies of the BioVU resource is described elsewhere^2^. ID traits were defined based on a hierarchical grouping of International Classification of Diseases, Ninth Revision (ICD-9) codes into phenotype codes (Phecode) representing clinical traits, as previously described^59,92^. (See below for a description of pathogen culture and viral test data in the BioVU individuals, including genera detected from different types of cultures.) We used version 1.2 of the Phecode Map containing 1,965 Phecodes based on 20,203 ICD-9 codes, which substantially improves signal-to-noise and more accurately reflects the clinical trait. Phecodes may exclude related phenotypes (e.g., in the case of Gram negative septicemia (Phecode = 038.1), the range of Phecodes given by 010-041.99, involving bacterial infection) and, importantly, include the definition of the appropriate control group^93^. Detailed description of Phecode trait maps can be found at phewascatalog.org. As an efficient and viable model for human genetics research, the Phecode system has been used to perform phenome-wide association studies (PheWAS) for validation of known genetic associations and discovery of new genetic disorders^59,60^.

### Pathogen culture and virology data linked to whole-genome genetic information

For individuals with whole-genome genetic information, we analyzed pathogen (bacterial, mycobacterial, and fungal) culture data derived from the following positive cultures for the indicated clinical samples: 1) blood (n = 7,699), 2) sputum (n = 2,478), 3) sinus/nasopharyngeal (n = 1,820), 4) bronchial-alveolar lavage (n = 1,265), and 5) tracheal sampling (n = 422). Furthermore, we analyzed a respiratory panel containing 28 viral strains from 2,890 individuals with whole-genome genetic information. Viral strains included the following: 1) Adenovirus, 2) Bocavirus, 3) Bordetella parapertussis, 4) Bordetella pertussis, 5) Chlamydia pneumoniae, 6) Coronavirus 229E, 7) Coronavirus HKU1, 8) Coronavirus NL63, 9) Coronavirus NOS, 10) Coronavirus OC43, 11) Enterovirus/Rhinovirus, 12) Human Metapneumovirus, 13) Influenza A, 14) Influenza A, H1, 15) Influenza A, H1N1, 16) Influenza A, H3, 17) Influenza B, 18) Mycoplasma pneumoniae, 19) Parainfluenza, 20) Parainfluenza 1, 21) Parainfluenza 2, 22) Parainfluenza 3, 23) Parainfluenza 4, 24) Respiratory syncytial virus (RSV), 25) RSV, A, 26) RSV, B, and 27) Rhinovirus. The pathogen information for each individual in our study included: 1) Total number of cultures; 2) Number of negative cultures (i.e., no pathogen growth); 3) Number of ambiguous cultures (i.e., normal upper respiratory bacteria or low level contamination); 4) Number of positive cultures (i.e., the number of cultures with growth consistent with clinical infection); 5) Genus or genera isolated (up to 96 unique genera per sample site), which ranged from zero to 10 per sample.

### GWAS of ID traits

GWAS of the ID traits were performed on the 23,294 and 4,321 BioVU individuals of European and African ancestry, respectively. Quality control pre-processing and SNP-level imputation were conducted, as previously described^60^. Genomic ancestry was quantified using principal components analysis of the genotype data^94,95^. GWAS of the ID traits were conducted in separate European and African genomic ancestry cohorts. Within each genomic ancestry, the association analysis was performed using age, gender, batch, and the first five principal components as covariates.

### Conditional SNP-level analysis

We performed conditional analysis on the top GWAS association with the ID trait (in this case, bacterial pneumonia) to determine whether it was driven by a related covariate (in this case, cystic fibrosis status). We used logistic regression to model the conditional probability of the infectious disease:

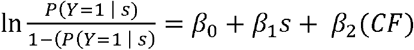

where *s* is the genotype at the sentinel variant, *Y* is the disease (i.e., bacterial pneumonia) status, and *CF* is the covariate of interest (i.e., cystic fibrosis).

### Transcriptome-wide association studies (TWAS) using PrediXcan

We performed multi-tissue PrediXcan^7,14,15^ in the 23,294 and 4,321 BioVU individuals of European and African ancestry, respectively. As in the GWAS analyses, these were conducted in the separate genomic ancestry cohorts. Experiment-wide significance (adjusted p < 0.05) was determined using Bonferroni correction for the total number of genes tested (n = 9,868) across 35 phenotypes (i.e., p < 1.4×10^−7^). Trait-specific significance (adjusted p < 0.05) was determined using Bonferroni correction for the total number of genes tested (n = 9,868, p < 5.07×10^−6^). Genomic ancestry was quantified using principal components analysis^94,95^. TWAS results were visualized using PhenoGram^96^.

### GWAS and TWAS Replication

For each ID trait, replication of GWAS and TWAS was performed using the cohort with the largest sample size (among BioVU^2^, the UK Biobank^1^, and FinnGen consortia^16^) as discovery and the remaining biobank(s) as replication. We used the UK Biobank (http://www.nealelab.is/uk-biobank) and the FinnGen (https://www.finngen.fi/en/access_results) summary results to generate the gene-level associations. GTEx v6p models (in 44 tissues) were used to generate tissue-level results.

### Classification of pathogen infection based on serology and culture data using several classifiers

Let *x* be a classifier (e.g., the Phecode or a logistic regression classifier) of serology and culture data based infection for a given pathogen, with probability density *φ* _+_(*x*) for positive instances and probability density *φ*_−_(*x*) for negative instances. The ROC curve plots the specificity (SP) and sensitivity (SN) at various thresholds:

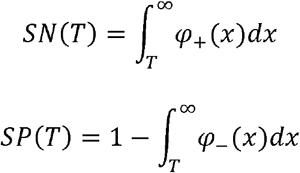

The area Ω under the curve (AUC) is given by:

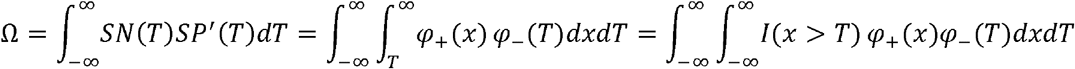

where *I*(*A*) is the indicator function, i.e., equal to one if (*x,T*) ∈*A* and zero otherwise. The last equals the probability that the classifier *X* ranks a randomly chosen positive instance (of culture data based infection) higher than a randomly chosen negative instance. We note that the expression for Ω suggests other metrics of interest, for example:

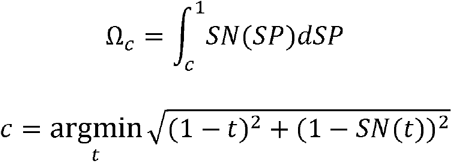

Here (*c,SN*(*c*) is the point on the ROC curve closest to true positive rate of 1 and false positive rate of 0. We estimated the sampling distribution of Ω (including standard error), using bootstrapping (n = 100) ^97^. We used the pROC package for visualization.

### Leveraging GWAS of human microbiome traits to extend GWAS of ID traits

We leveraged genome-wide associations of microbiome traits, involving 155 pathogens derived from phylogenetic analysis of 16S rRNA gene sequences^17^.

### Causal inference by Mendelian Randomization

To infer causality between the infectious diseases and potential complications, we performed Mendelian Randomization (MR^18,21^) in 23,294 individuals of European ancestry in BioVU. To define instrumental variables (IVs), we clumped the exposure-associated SNPs with high linkage disequilibrium (LD) using Plink1.9 (p < 1×10^−5^, r^2^ = 0.01). Only biallelic non-palindromic variants were considered as IVs. Considering the pervasive horizontal pleiotropy in human genetic variation^98^, we applied summary statistics based MR-Egger regression^65^. MR-Egger regression generalizes the inverse-variance weighted method, where the intercept is assumed to be zero. We also used the weighted-median estimator^66^ to test the causal effect of the exposure trait on the outcome. We leveraged the R package ‘MendelianRandomization’.

### High-throughput Human in vitrO Susceptibility Testing (Hi-HOST)

We generated an atlas of TWAS associations with 79 pathogen-induced cellular traits – including infectivity and replication, cytokine levels, and host cell death^19^ using the Hi-HOST platform^99,100^. A list of populations, pathogens and project description may be found at http://h2p2.oit.duke.edu/About/, and phenotype definitions and family-based GWAS of the Hi-HOST Phenome Project were previously described^19^. Briefly, lymphoblastoid cell lines (LCLs) from the 1000 Genomes Consortium^101^ were obtained from the Coriell Institute. The LCLs represented diverse populations, including ESN (Esan in Nigeria), GWD (Gambians in Western Divisions in the Gambia), IBS (Iberian Population in Spain), and KHV (Kinh in Ho Chi Minh City, Vietnam). LCLs were cultured in RPMI 1640 media containing 10% fetal bovine serum, 2 mM glutamine, 100 U/ml of penicillin-G, and 100 mg/ml streptomycin for 8 days prior to experimental use, as previously described^19^. *Chlamydia trachomatis* infection of LCLs was performed using *C. trachomatis* LGV-L2 Rif^R^ pGFP::SW2^102^. *Salmonella* infection was performed using pMMB67GFP^103^, and sifA deletion was constructed using lambda red and validated using PCR^100,104^. *Candida albicans* SC5314 infection was performed as previously described^105^ and levels of fibroblast growth factor 2 were measured using enzyme linked immunosorbent assays. *Staphylococcus aureus* toxin (alpha-hemolysin) was obtained from Sigma and applied to LCLs at a concentration of 1 μg/ml for 23 hours. Cell death was measured using 7-AAD staining and flow cytometry. Additional experimental details can be found at http://h2p2.oit.duke.edu/About/.

We estimated the gene-level effect size on the Hi-HOST phenotypes, using GWAS summary statistics^106^ in each of the 44 GTEx tissues (version 6p)^107^. The gene expression prediction model was trained using GTEx as the reference dataset (https://zenodo.org/record/3572842/files/GTEx-V6p-HapMap-2016-09-08.tar.gz). The gene-level effect size was estimated using S-PrediXcan after allele harmonization^106^. We also applied MultiXcan to improve the ability to identify potential target genes^15^. In brief, MultiXcan regresses the cellular trait on the principal components of the predicted expression data across all the available tissues. For each gene, MultiXcan yields a joint effect estimate across the 44 tissues. We applied the summary-statistic based version (S-MultiXcan) and followed the guides from the tool’s webpage https://github.com/hakyimlab/MetaXcan.

### Data and software availability

All code is available at the project’s GitHub page: https://github.com/gamazonlab/infectiousDiseaseResource. Further information and requests for resources and reagents should be directed to and will be fulfilled by the Corresponding Author, Eric R. Gamazon.

## Data Availability

All results and code are available at the links below.

https://github.com/gamazonlab/infectiousDiseaseResource

## AUTHOR CONTRIBUTIONS

A.T.H. and E.R.G. conceived and designed the study. A.T.H., D.Z., R.L.S., L.B., S.J.S., D.C.K., and E.R.G. contributed new methodologies. A.T.H., D.Z., R.L.S., L.B., L.W., S.S.Z., S.J.S., D.C.K., and E.R.G acquired, processed, and analyzed data. A.T.H. and E.R.G. drafted the manuscript. All authors critically revised the manuscript for important intellectual content.

## ACKNOWLEDGEMENTS

A.T.H. is supported by the National Institutes of Health (F30HL143826) and Vanderbilt University Medical Scientist Training Program (T32GM007347). E.R.G. is supported by the National Human Genome Research Institute of the National Institutes of Health under Award Numbers R35HG010718 and R01HG011138. E.R.G. and S.S.Z. are funded by the National Heart, Lung, & Blood Institute of the National Institutes of Health under Award Number R01HL133559. The content is solely the responsibility of the authors and does not necessarily represent the official views of the National Institutes of Health. E.R.G. has also significantly benefitted from a Fellowship at Clare Hall, University of Cambridge (UK) and is grateful to the President and Fellows of the college for a stimulating intellectual home. Genomic data are also supported by individual investigator-led projects including U01-HG004798, R01-NS032830, RC2-GM092618, P50-GM115305, U01-HG006378, U19-HL065962, and R01-HD074711. Additional funding sources for BioVU are listed at https://victr.vanderbilt.edu/pub/biovu/. L.B. is supported by R01-LM010685. S.J.S. is supported by an NIH Director’s Pioneer and Transformative Awards DP1-HD086071 and R01-AI145057. D.C.K. is supported by R01-AI118903, R21-AI144586, and R21-AI146520. D.C.K. and L.W. are supported by R21-AI133305.

## COMPETING INTERESTS

E.R.G. receives an honorarium from the journal *Circulation Research* of the American Heart Association, as a member of the Editorial Board. The other authors declare no competing interests.

## Notes

### Author Declarations

Vanderbilt University Medical Center IRB

## REFERENCES

1 Bycroft, C. et al. The UK Biobank resource with deep phenotyping and genomic data. Nature 562, 203–209, doi:10.1038/s41586-018-0579-z (2018).

2 Roden, D. M. et al. Development of a large-scale de-identified DNA biobank to enable personalized medicine. Clinical pharmacology and therapeutics 84, 362–369, doi:10.1038/clpt.2008.89 (2008).

3 Cotsapas, C. et al. Pervasive sharing of genetic effects in autoimmune disease. PLoS Genet 7, e1002254, doi:10.1371/journal.pgen.1002254 (2011).

4 Gamazon, E. R., Zwinderman, A. H., Cox, N. J., Denys, D. & Derks, E. M. Multi-tissue transcriptome analyses identify genetic mechanisms underlying neuropsychiatric traits. Nat Genet 51, 933–940, doi:10.1038/s41588-019-0409-8 (2019).

5 Watanabe, K. et al. A global overview of pleiotropy and genetic architecture in complex traits. Nat Genet, doi:10.1038/s41588-019-0481-0 (2019).

6 Bulik-Sullivan, B. et al. An atlas of genetic correlations across human diseases and traits. Nat Genet 47, 1236–1241, doi:10.1038/ng.3406 (2015).

7 Gamazon, E. R. et al. Using an atlas of gene regulation across 44 human tissues to inform complex disease- and trait-associated variation. Nat Genet 50, 956–967, doi:10.1038/s41588-018-0154-4 (2018).

8 Shi, H., Kichaev, G. & Pasaniuc, B. Contrasting the Genetic Architecture of 30 Complex Traits from Summary Association Data. Am J Hum Genet 99, 139–153, doi:10.1016/j.ajhg.2016.05.013 (2016).

9 Chapman, S. J. & Hill, A. V. Human genetic susceptibility to infectious disease. Nat Rev Genet 13, 175–188, doi:10.1038/nrg3114 (2012).

10 de Bakker, P. I. & Telenti, A. Infectious diseases not immune to genome-wide association. Nat Genet 42, 731–732, doi:10.1038/ng0910-731 (2010).

11 Hill, A. V. The genomics and genetics of human infectious disease susceptibility. Annu Rev Genomics Hum Genet 2, 373–400, doi:10.1146/annurev.genom.2.1.373 (2001).

12 Ko, D. C. & Urban, T. J. Understanding human variation in infectious disease susceptibility through clinical and cellular GWAS. PLoS Pathog 9, e1003424, doi:10.1371/journal.ppat.1003424 (2013).

13 van de Vosse, E., van Dissel, J. T. & Ottenhoff, T. H. Genetic deficiencies of innate immune signalling in human infectious disease. Lancet Infect Dis 9, 688–698, doi:10.1016/s1473-3099(09)70255-5 (2009).

14 Gamazon, E. R. et al. A gene-based association method for mapping traits using reference transcriptome data. Nat Genet 47, 1091–1098, doi:10.1038/ng.3367 (2015).

15 Barbeira, A. N. et al. Integrating predicted transcriptome from multiple tissues improves association detection. PLoS Genet 15, e1007889, doi:10.1371/journal.pgen.1007889 (2019).

16 Locke, A. E. et al. Exome sequencing of Finnish isolates enhances rare-variant association power. Nature 572, 323–328, doi:10.1038/s41586-019-1457-z (2019).

17 Hughes, D. A. et al. Genome-wide associations of human gut microbiome variation and implications for causal inference analyses. Nature microbiology 5, 1079–1087, doi:10.1038/s41564-020-0743-8 (2020).

18 Lawlor, D. A., Harbord, R. M., Sterne, J. A., Timpson, N. & Davey Smith, G. Mendelian randomization: using genes as instruments for making causal inferences in epidemiology. Stat Med 27, 1133–1163, doi:10.1002/sim.3034 (2008).

19 Wang, L. et al. An Atlas of Genetic Variation Linking Pathogen-Induced Cellular Traits to Human Disease. Cell Host Microbe 24, 308-323.e306, doi:10.1016/j.chom.2018.07.007 (2018).

20 Gusev, A. et al. Integrative approaches for large-scale transcriptome-wide association studies. Nature genetics 48, 245–252, doi:10.1038/ng.3506 (2016).

21 Davey Smith, G. & Hemani, G. Mendelian randomization: genetic anchors for causal inference in epidemiological studies. Human molecular genetics 23, R89–98, doi:10.1093/hmg/ddu328 (2014).

22 García-Montero, M. et al. Pneumonia caused by Listeria monocytogenes. Respiration 62, 107–109, doi:10.1159/000196402 (1995).

23 Lyczak, J. B., Cannon, C. L. & Pier, G. B. Lung infections associated with cystic fibrosis. Clin Microbiol Rev 15, 194–222, doi:10.1128/cmr.15.2.194-222.2002 (2002).

24 Balsa, E. et al. NDUFA4 is a subunit of complex IV of the mammalian electron transport chain. Cell Metab 16, 378–386, doi:10.1016/j.cmet.2012.07.015 (2012).

25 Tartey, S. et al. Essential Function for the Nuclear Protein Akirin2 in B Cell Activation and Humoral Immune Responses. J Immunol 195, 519–527, doi:10.4049/jimmunol.1500373 (2015).

26 Tartey, S. et al. Akirin2 is critical for inducing inflammatory genes by bridging IkappaB-zeta and the SWI/SNF complex. Embo j 33, 2332–2348, doi:10.15252/embj.201488447 (2014).

27 Revill, K. et al. Genome-wide methylation analysis and epigenetic unmasking identify tumor suppressor genes in hepatocellular carcinoma. Gastroenterology 145, 1424-1435.e1421-1425, doi:10.1053/j.gastro.2013.08.055 (2013).

28 Sun, W., Li, J., Jiang, H. G., Ge, L. P. & Wang, Y. Diagnostic value of MUC1 and EpCAM mRNA as tumor markers in differentiating benign from malignant pleural effusion. QJM : monthly journal of the Association of Physicians 107, 1001–1007, doi:10.1093/qjmed/hcu130 (2014).

29 Power, R. A., Parkhill, J. & de Oliveira, T. Microbial genome-wide association studies: lessons from human GWAS. Nature Reviews Genetics 18, 41–50, doi:10.1038/nrg.2016.132 (2017).

30 Domingo, P. et al. Spontaneous rupture of the spleen associated with pneumonia. European journal of clinical microbiology & infectious diseases : official publication of the European Society of Clinical Microbiology 15, 733–736, doi:10.1007/bf01691960 (1996).

31 Gerstein, A. R., Riegel, N. & Dennis, M. Ruptured Spleen Simulating Pneumonia. JAMA 199, 589–589, doi:10.1001/jama.1967.03120080123033 (1967).

32 Huang da, W., Sherman, B. T. & Lempicki, R. A. Systematic and integrative analysis of large gene lists using DAVID bioinformatics resources. Nat Protoc 4, 44–57, doi:10.1038/nprot.2008.211 (2009).

33 Soderholm, S. et al. Phosphoproteomics to Characterize Host Response During Influenza A Virus Infection of Human Macrophages. Molecular & cellular proteomics : MCP 15, 3203–3219, doi:10.1074/mcp.M116.057984 (2016).

34 Stahl, J. A. et al. Phosphoproteomic analyses reveal signaling pathways that facilitate lytic gammaherpesvirus replication. PLoS Pathog 9, e1003583, doi:10.1371/journal.ppat.1003583 (2013).

35 Murray, L. A., Sheng, X. & Cristea, I. M. Orchestration of protein acetylation as a toggle for cellular defense and virus replication. Nat Commun 9, 4967, doi:10.1038/s41467-018-07179-w (2018).

36 Taylor, M. P., Koyuncu, O. O. & Enquist, L. W. Subversion of the actin cytoskeleton during viral infection. Nat Rev Microbiol 9, 427–439, doi:10.1038/nrmicro2574 (2011).

37 Aktories, K. & Barbieri, J. T. Bacterial cytotoxins: targeting eukaryotic switches. Nat Rev Microbiol 3, 397–410, doi:10.1038/nrmicro1150 (2005).

38 Yu, B., Cheng, H. C., Brautigam, C. A., Tomchick, D. R. & Rosen, M. K. Mechanism of actin filament nucleation by the bacterial effector VopL. Nat Struct Mol Biol 18, 1068–1074, doi:10.1038/nsmb.2110 (2011).

39 Zahm, J. A. et al. The bacterial effector VopL organizes actin into filament-like structures. Cell 155, 423–434, doi:10.1016/j.cell.2013.09.019 (2013).

40 Xie, X. et al. Systematic discovery of regulatory motifs in human promoters and 3’ UTRs by comparison of several mammals. Nature 434, 338–345, doi:10.1038/nature03441 (2005).

41 Ammari, M. G., Gresham, C. R., McCarthy, F. M. & Nanduri, B. HPIDB 2.0: a curated database for host-pathogen interactions. Database : the journal of biological databases and curation 2016, doi:10.1093/database/baw103 (2016).

42 Na, Y. R. et al. The early synthesis of p35 and activation of CDK5 in LPS-stimulated macrophages suppresses interleukin-10 production. Science signaling 8, ra121–ra121, doi:10.1126/scisignal.aab3156 (2015).

43 Patrick, G. N. et al. Conversion of p35 to p25 deregulates Cdk5 activity and promotes neurodegeneration. Nature 402, 615–622, doi:10.1038/45159 (1999).

44 Tarricone, C. et al. Structure and regulation of the CDK5-p25(nck5a) complex. Molecular cell 8, 657–669 (2001).

45 Pfänder, P., Fidan, M., Burret, U., Lipinski, L. & Vettorazzi, S. Cdk5 Deletion Enhances the Anti-inflammatory Potential of GC-Mediated GR Activation During Inflammation. Frontiers in Immunology 10, doi:10.3389/fimmu.2019.01554 (2019).

46 Man, A., Slevin, M., Petcu, E. & Fraefel, C. The Cyclin-Dependent Kinase 5 Inhibitor Peptide Inhibits Herpes Simplex Virus Type 1 Replication. Scientific reports 9, 1260, doi:10.1038/s41598-018-37989-3 (2019).

47 Davis, Z. H. et al. Global mapping of herpesvirus-host protein complexes reveals a transcription strategy for late genes. Molecular cell 57, 349–360, doi:10.1016/j.molcel.2014.11.026 (2015).

48 Hay, A. J., Wolstenholme, A. J., Skehel, J. J. & Smith, M. H. The molecular basis of the specific anti-influenza action of amantadine. Embo j 4, 3021–3024 (1985).

49 Barnard, P., Payne, E. & McMillan, N. A. The human papillomavirus E7 protein is able to inhibit the antiviral and anti-growth functions of interferon-alpha. Virology 277, 411–419, doi:10.1006/viro.2000.0584 (2000).

50 Chellappan, S. et al. Adenovirus E1A, simian virus 40 tumor antigen, and human papillomavirus E7 protein share the capacity to disrupt the interaction between transcription factor E2F and the retinoblastoma gene product. Proc Natl Acad Sci U S A 89, 4549–4553, doi:10.1073/pnas.89.10.4549 (1992).

51 Phelps, W. C., Yee, C. L., Münger, K. & Howley, P. M. The human papillomavirus type 16 E7 gene encodes transactivation and transformation functions similar to those of adenovirus E1A. Cell 53, 539–547, doi:10.1016/0092-8674(88)90570-3 (1988).

52 Longworth, M. S. & Laimins, L. A. The binding of histone deacetylases and the integrity of zinc finger-like motifs of the E7 protein are essential for the life cycle of human papillomavirus type 31. J Virol 78, 3533–3541, doi:10.1128/jvi.78.7.3533-3541.2004 (2004).

53 Duro, D. et al. Activation of cyclin A gene expression by the cyclin encoded by human herpesvirus-8. J Gen Virol 80 (Pt 3), 549–555, doi:10.1099/0022-1317-80-3-549 (1999).

54 Ojala, P. M. et al. Kaposi’s sarcoma-associated herpesvirus-encoded v-cyclin triggers apoptosis in cells with high levels of cyclin-dependent kinase 6. Cancer Res 59, 4984–4989 (1999).

55 Ojala, P. M. et al. The apoptotic v-cyclin-CDK6 complex phosphorylates and inactivates Bcl-2. Nature cell biology 2, 819–825, doi:10.1038/35041064 (2000).

56 Goldstein, E. J., Citron, D. M., Peraino, V. A. & Cross, S. A. Desulfovibrio desulfuricans bacteremia and review of human Desulfovibrio infections. Journal of clinical microbiology 41, 2752–2754, doi:10.1128/jcm.41.6.2752-2754.2003 (2003).

57 Loubinoux, J., Bronowicki, J. P., Pereira, I. A., Mougenel, J. L. & Faou, A. E. Sulfate-reducing bacteria in human feces and their association with inflammatory bowel diseases. FEMS Microbiol Ecol 40, 107–112, doi:10.1111/j.1574-6941.2002.tb00942.x (2002).

58 Bastarache, L. et al. Phenotype risk scores identify patients with unrecognized Mendelian disease patterns. Science (New York, N.Y.) 359, 1233–1239, doi:10.1126/science.aal4043 (2018).

59 Denny, J. C. et al. Systematic comparison of phenome-wide association study of electronic medical record data and genome-wide association study data. Nature biotechnology 31, 1102–1110, doi:10.1038/nbt.2749 (2013).

60 Unlu, G. et al. Phenome-based approach identifies RIC1-linked Mendelian syndrome through zebrafish models, biobank associations and clinical studies. Nat Med 26, 98–109, doi:10.1038/s41591-019-0705-y (2020).

61 Hagiwara, K. et al. Mouse SWAM1 and SWAM2 are antibacterial proteins composed of a single whey acidic protein motif. J Immunol 170, 1973–1979, doi:10.4049/jimmunol.170.4.1973 (2003).

62 Niemöller, U. M. & Täuber, M. G. Brain edema and increased intracranial pressure in the pathophysiology of bacterial meningitis. European journal of clinical microbiology & infectious diseases : official publication of the European Society of Clinical Microbiology 8, 109–117, doi:10.1007/bf01963892 (1989).

63 Suetrong, B. & Walley, K. R. Lactic Acidosis in Sepsis: It’s Not All Anaerobic: Implications for Diagnosis and Management. Chest 149, 252–261, doi:10.1378/chest.15-1703 (2016).

64 Isaac, R. et al. TM7SF3, a novel p53-regulated homeostatic factor, attenuates cellular stress and the subsequent induction of the unfolded protein response. Cell Death Differ 24, 132–143, doi:10.1038/cdd.2016.108 (2017).

65 Bowden, J., Davey Smith, G. & Burgess, S. Mendelian randomization with invalid instruments: effect estimation and bias detection through Egger regression. Int J Epidemiol 44, 512–525, doi:10.1093/ije/dyv080 (2015).

66 Bowden, J., Davey Smith, G., Haycock, P. C. & Burgess, S. Consistent Estimation in Mendelian Randomization with Some Invalid Instruments Using a Weighted Median Estimator. Genetic epidemiology 40, 304–314, doi:10.1002/gepi.21965 (2016).

67 Zheng, T. et al. IL-13 receptor alpha2 selectively inhibits IL-13-induced responses in the murine lung. J Immunol 180, 522–529, doi:10.4049/jimmunol.180.1.522 (2008).

68 Gómez-Moreno, J., Moar, C., Román, F., Pérez-Maestu, R. & López de Letona, J. M. Salmonella endocarditis presenting as cerebral hemorrhage. Eur J Intern Med 11, 96–97, doi:10.1016/s0953-6205(00)00060-1 (2000).

69 Tian, C. et al. Genome-wide association and HLA region fine-mapping studies identify susceptibility loci for multiple common infections. Nat Commun 8, 599, doi:10.1038/s41467-017-00257-5 (2017).

70 Casanova, J. L. Human genetic basis of interindividual variability in the course of infection. Proc Natl Acad Sci U S A 112, E7118–7127, doi:10.1073/pnas.1521644112 (2015).

71 Casanova, J. L. Severe infectious diseases of childhood as monogenic inborn errors of immunity. Proc Natl Acad Sci U S A 112, E7128–7137, doi:10.1073/pnas.1521651112 (2015).

72 So, H. C. et al. Analysis of genome-wide association data highlights candidates for drug repositioning in psychiatry. Nat Neurosci 20, 1342–1349, doi:10.1038/nn.4618 (2017).

73 El-Bacha, T. & Da Poian, A. T. Virus-induced changes in mitochondrial bioenergetics as potential targets for therapy. The international journal of biochemistry & cell biology 45, 41–46, doi:10.1016/j.biocel.2012.09.021 (2013).

74 Labbé, K. & Saleh, M. Cell death in the host response to infection. Cell Death Differ 15, 1339–1349, doi:10.1038/cdd.2008.91 (2008).

75 Brouwer, W. P. et al. Genome Wide Association Study Identifies Genetic Variants Associated With Early And Sustained Response To (Peg)Interferon In Chronic Hepatitis B Patients: The GIANT-B Study. Clinical infectious diseases : an official publication of the Infectious Diseases Society of America, doi:10.1093/cid/ciz084 (2019).

76 Liang, X. et al. Macrophage FABP4 is required for neutrophil recruitment and bacterial clearance in Pseudomonas aeruginosa pneumonia. Faseb j 33, 3562–3574, doi:10.1096/fj.201802002R (2019).

77 Pan, Y. et al. Survival of tissue-resident memory T cells requires exogenous lipid uptake and metabolism. Nature 543, 252–256, doi:10.1038/nature21379 (2017).

78 Saitoh, T. et al. Atg9a controls dsDNA-driven dynamic translocation of STING and the innate immune response. Proc Natl Acad Sci U S A 106, 20842–20846, doi:10.1073/pnas.0911267106 (2009).

79 Sharfe, N., Dadi, H. K., Shahar, M. & Roifman, C. M. Human immune disorder arising from mutation of the alpha chain of the interleukin-2 receptor. Proc Natl Acad Sci U S A 94, 3168–3171, doi:10.1073/pnas.94.7.3168 (1997).

80 Tsuboi, S. & Meerloo, J. Wiskott-Aldrich syndrome protein is a key regulator of the phagocytic cup formation in macrophages. The Journal of biological chemistry 282, 34194–34203, doi:10.1074/jbc.M705999200 (2007).

81 Walenna, N. F. et al. Chlamydia pneumoniae exploits adipocyte lipid chaperone FABP4 to facilitate fat mobilization and intracellular growth in murine adipocytes. Biochem Biophys Res Commun 495, 353–359, doi:10.1016/j.bbrc.2017.11.005 (2018).

82 Willis, K. L., Patel, S., Xiang, Y. & Shisler, J. L. The effect of the vaccinia K1 protein on the PKR-eIF2alpha pathway in RK13 and HeLa cells. Virology 394, 73–81, doi:10.1016/j.virol.2009.08.020 (2009).

83 Yu, Z. et al. USP1-UAF1 deubiquitinase complex stabilizes TBK1 and enhances antiviral responses. The Journal of experimental medicine 214, 3553–3563, doi:10.1084/jem.20170180 (2017).

84 Zhang, X. et al. Human intracellular ISG15 prevents interferon-alpha/beta over-amplification and auto-inflammation. Nature 517, 89–93, doi:10.1038/nature13801 (2015).

85 Lanzi, G. et al. A novel primary human immunodeficiency due to deficiency in the WASP-interacting protein WIP. The Journal of experimental medicine 209, 29–34, doi:10.1084/jem.20110896 (2012).

86 Herman, M. et al. Heterozygous TBK1 mutations impair TLR3 immunity and underlie herpes simplex encephalitis of childhood. The Journal of experimental medicine 209, 1567–1582, doi:10.1084/jem.20111316 (2012).

87 Bouhaddou, M. et al. The Global Phosphorylation Landscape of SARS-CoV-2 Infection. Cell, doi:https://doi.org/10.1016/j.cell.2020.06.034(2020).

88 Goodrich, J. K. et al. Genetic Determinants of the Gut Microbiome in UK Twins. Cell Host Microbe 19, 731–743, doi:10.1016/j.chom.2016.04.017 (2016).

89 Hall, A. B., Tolonen, A. C. & Xavier, R. J. Human genetic variation and the gut microbiome in disease. Nat Rev Genet 18, 690–699, doi:10.1038/nrg.2017.63 (2017).

90 Kurilshikov, A., Wijmenga, C., Fu, J. & Zhernakova, A. Host Genetics and Gut Microbiome: Challenges and Perspectives. Trends in immunology 38, 633–647, doi:10.1016/j.it.2017.06.003 (2017).

91 Taylor, A. E. et al. Investigating the possible causal association of smoking with depression and anxiety using Mendelian randomisation meta-analysis: the CARTA consortium. BMJ Open 4, e006141, doi:10.1136/bmjopen-2014-006141 (2014).

92 Denny, J. C. et al. PheWAS: demonstrating the feasibility of a phenome-wide scan to discover gene-disease associations. Bioinformatics 26, 1205–1210, doi:10.1093/bioinformatics/btq126 (2010).

93 Wei, W.-Q. et al. Evaluating phecodes, clinical classification software, and ICD-9-CM codes for phenome-wide association studies in the electronic health record. PloS one 12, e0175508–e0175508, doi:10.1371/journal.pone.0175508 (2017).

94 Derks, E. M., Zwinderman, A. H. & Gamazon, E. R. The Relation Between Inflation in Type-I and Type-II Error Rate and Population Divergence in Genome-Wide Association Analysis of Multi-Ethnic Populations. Behavior genetics 47, 360–368, doi:10.1007/s10519-017-9837-3 (2017).

95 Price, A. L. et al. Principal components analysis corrects for stratification in genome-wide association studies. Nat Genet 38, 904–909, doi:10.1038/ng1847 (2006).

96 Wolfe, D., Dudek, S., Ritchie, M. & Pendergrass, S. Visualizing genomic information across chromosomes with PhenoGram. BioData Mining 6, 18 – 18 (2013).

97 Efron, B. Bootstrap Methods: Another Look at the Jackknife. Ann. Statist. 7, 1–26, doi:10.1214/aos/1176344552 (1979).

98 Jordan, D. M., Verbanck, M. & Do, R. HOPS: a quantitative score reveals pervasive horizontal pleiotropy in human genetic variation is driven by extreme polygenicity of human traits and diseases. Genome Biol 20, 222, doi:10.1186/s13059-019-1844-7 (2019).

99 Ko, D. C. et al. Functional genetic screen of human diversity reveals that a methionine salvage enzyme regulates inflammatory cell death. Proc Natl Acad Sci U S A 109, E2343–2352, doi:10.1073/pnas.1206701109 (2012).

100 Ko, D. C. et al. A genome-wide in vitro bacterial-infection screen reveals human variation in the host response associated with inflammatory disease. Am J Hum Genet 85, 214–227, doi:10.1016/j.ajhg.2009.07.012 (2009).

101 Auton, A. et al. A global reference for human genetic variation. Nature 526, 68–74, doi:10.1038/nature15393 (2015).

102 Saka, H. A. et al. Quantitative proteomics reveals metabolic and pathogenic properties of Chlamydia trachomatis developmental forms. Molecular microbiology 82, 1185–1203, doi:10.1111/j.1365-2958.2011.07877.x (2011).

103 Pujol, C. & Bliska, J. B. The ability to replicate in macrophages is conserved between Yersinia pestis and Yersinia pseudotuberculosis. Infect Immun 71, 5892–5899, doi:10.1128/iai.71.10.5892-5899.2003 (2003).

104 Datsenko, K. A. & Wanner, B. L. One-step inactivation of chromosomal genes in Escherichia coli K-12 using PCR products. Proc Natl Acad Sci U S A 97, 6640–6645, doi:10.1073/pnas.120163297 (2000).

105 Odds, F. C., Brown, A. J. & Gow, N. A. Candida albicans genome sequence: a platform for genomics in the absence of genetics. Genome Biol 5, 230, doi:10.1186/gb-2004-5-7-230 (2004).

106 Barbeira, A. N. et al. Exploring the phenotypic consequences of tissue specific gene expression variation inferred from GWAS summary statistics. Nat Commun 9, 1825, doi:10.1038/s41467-018-03621-1 (2018).

107 Battle, A., Brown, C. D., Engelhardt, B. E. & Montgomery, S. B. Genetic effects on gene expression across human tissues. Nature 550, 204–213, doi:10.1038/nature24277 (2017).

