## Supplementary Information for "The genetic architecture of human infectious diseases and pathogen-induced cellular phenotypes"

**SUPPLEMENTAL INFORMATION**

**Figure S1.** Replication of ID trait TWAS in the UK Biobank. (A) Q-Q plot of UK Biobank replication p-values for the gene-level associations with bacterial pneumonia in lung tissue. (B) Q-Q plot of UK Biobank replication p-values for the gene-level associations with meningitis in hippocampus. (C) Q-Q plot of UK Biobank replication p-values for the gene-level associations with meningitis in cerebellar hemisphere. (D) Q-Q plot of UK Biobank replication p-values for the gene-level associations with meningitis in hypothalamus. (E) Q-Q plot of UK Biobank replication p-values for the gene-level associations with encephalitis in caudate. (F) Q-Q plot of UK Biobank replication p-values for the gene-level associations with encephalitis in cortex. (G) Q-Q plot of UK Biobank replication p-values for the gene-level associations with encephalitis in cerebellum.

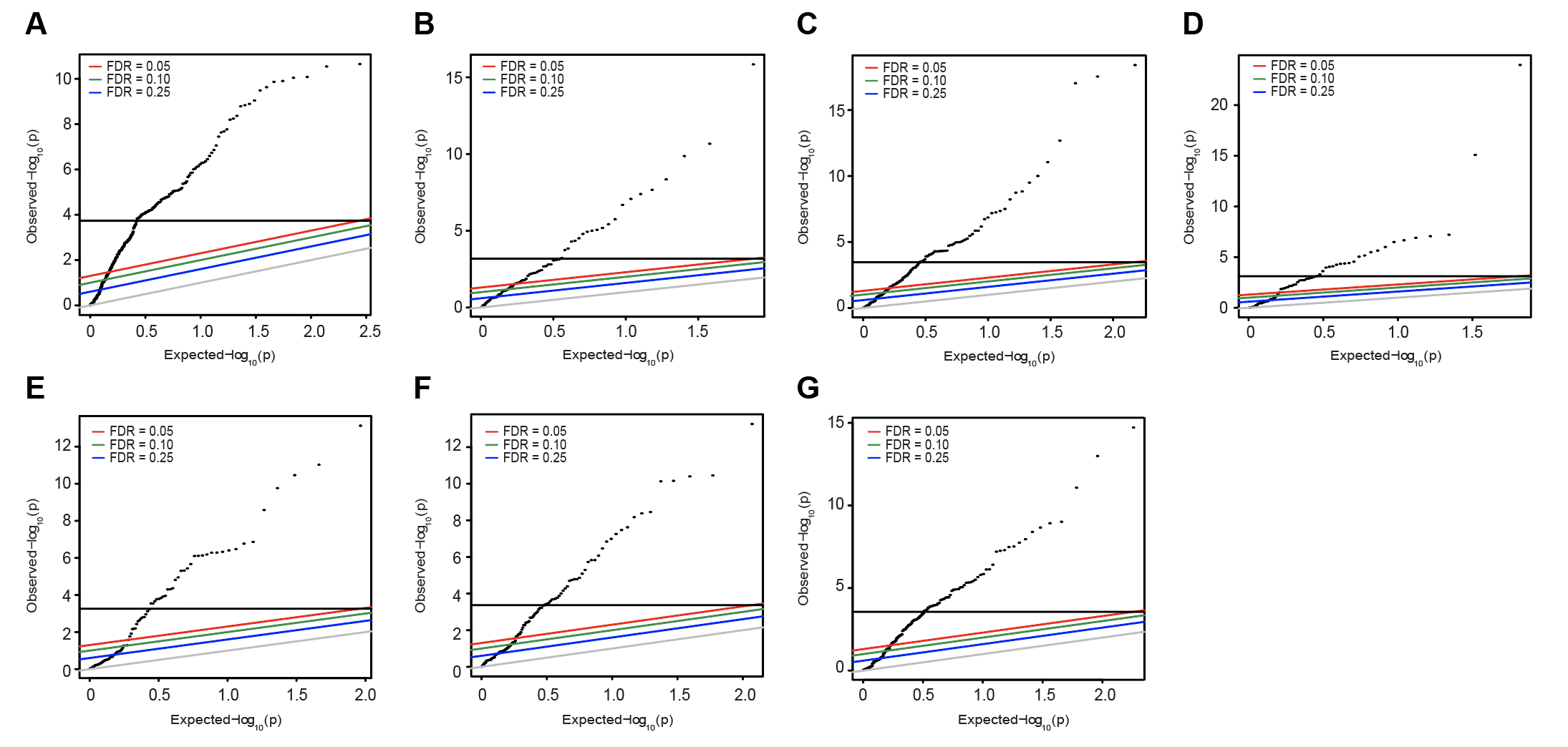

**Figure S2.** Expression profile of ID-associated genes in human tissues is consistent with affected tissues, puts constraints on diagnostic utility of a surrogate tissue, and shows the diversity of potential mechanisms for clinical complications. (A) ID-associated genes are more likely to be ubiquitously expressed compared to complement genes (Mann Whitney U test, p= 7.5x10^-4^), in part reflecting the statistically powerful cross-tissue TWAS approach we implemented to identify ID associations. The histogram shows the empirical distribution of tissue specificity, as quantified by the tau statistic (x-axis). (B) *NDUFA* expression across tissues available in GTEx. The gene is the most significant association (with intestinal infection) in our study and is ubiquitously expressed throughout the alimentary canal but displays considerably lower expression in whole blood, which may limit the utility of the tissue for diagnostic purposes. (C) *TOR4A* expression across tissues available in GTEx. The gene, which is associated with bacterial pneumonia in BioVU and is replicated in UK Biobank, is most abundantly expressed in lung, consistent with the tissue of pathology, but also in spleen, which is ruptured as a lethal complication of the infection. Adverse outcomes and complications may therefore arise in part from the effect of a susceptibility gene in a secondary tissue, i.e., distinct from the tissue of pathology for the infection.

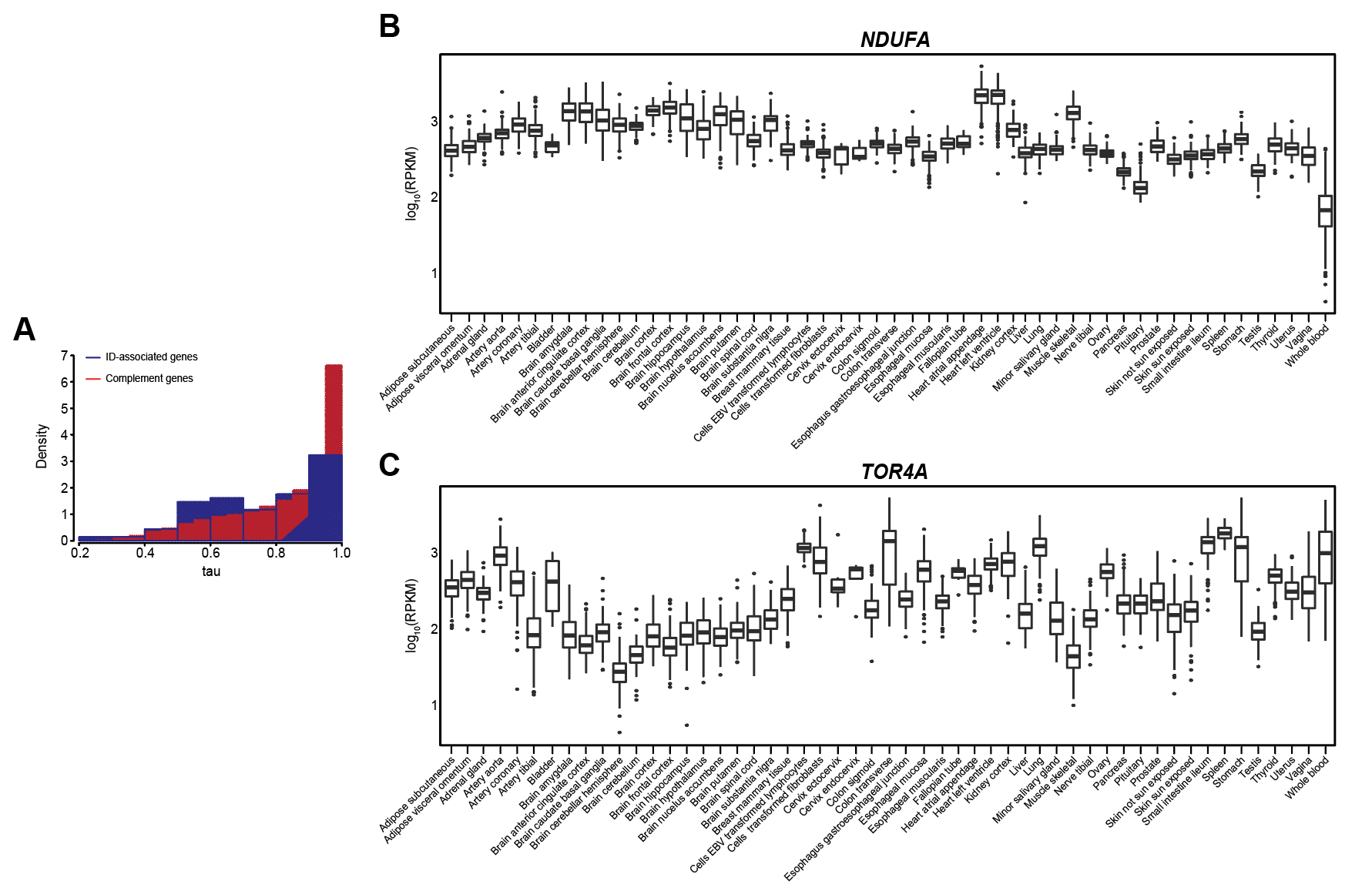

**Figure S3. Bacterial, fungal, and viral genus identification from clinical samples linked to whole-genome information reveals insights into host colonization and infection.** (A) bronco-alveolar lavage cultures (n = 1,854 positive pathogen cultures across 48 genera) from 1,265 individuals; (B) sputum cultures (n = 3,289 positive pathogen cultures across 57 genera) from 2,478 individuals; (C) sinus/nasopharyngeal cultures (n = 2,077 positive pathogen cultures across 39 genera) from 1,820 individuals; (D) tracheal cultures (n = 488 positive pathogen cultures across 19 genera) from 422 individuals. (E). Viruses identified across 64,327 clinical samples for 27 viral genera isolated from 2,890 individuals.

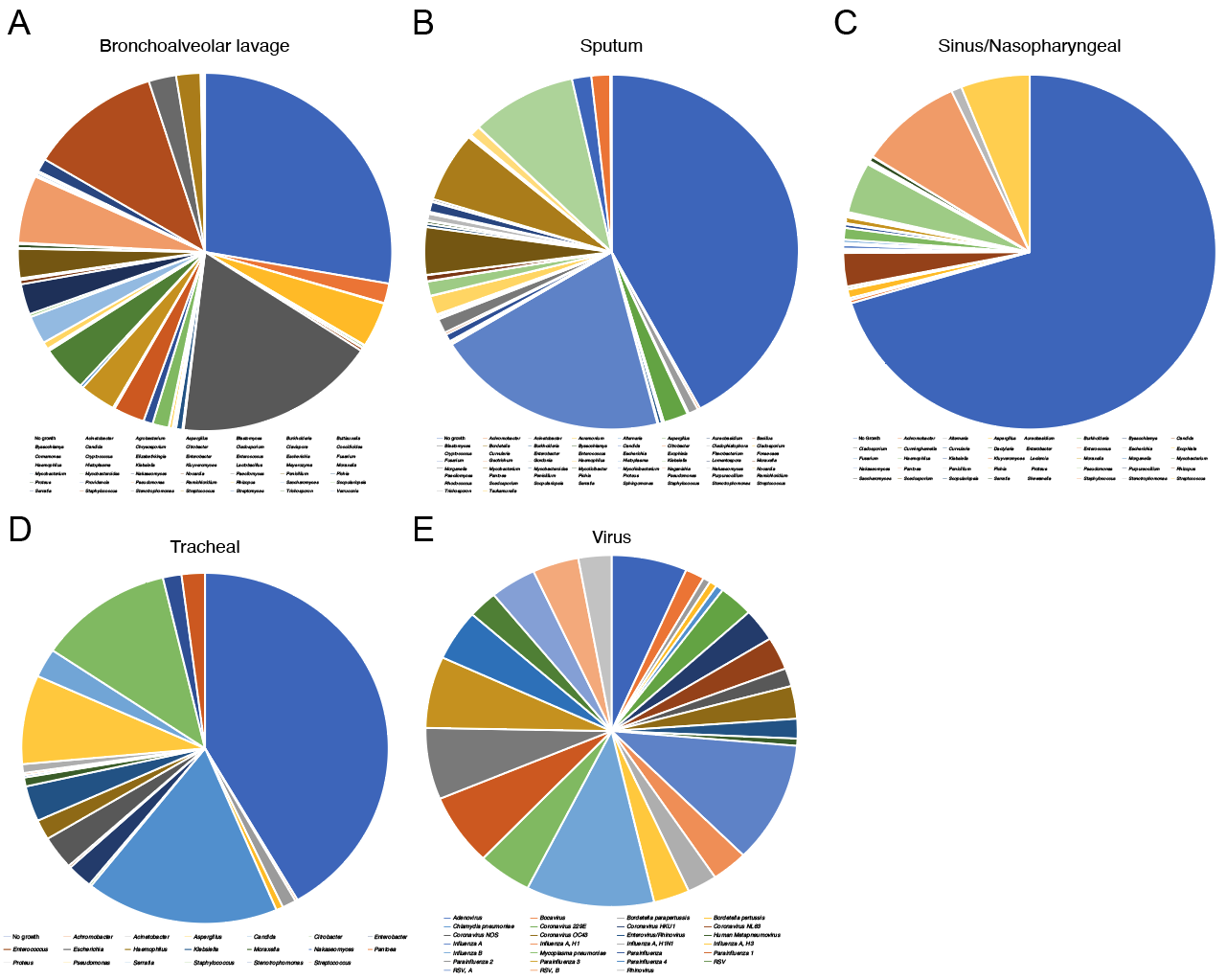

**Table S1.** Significant trait-specific SNP-level GWAS associations with individual infectious disease phenotypes (for which number of cases > 100). All variants listed reached genome wide significance (p < 5.0x10^-8^).

| **Phenotype** | **Phecode** | **SNP** | **OR** | **P value** |
| --- | --- | --- | --- | --- |
| Bacterial pneumonia | 480.1 | rs17139584 | 2.85 | 1.21x10^-36^ |
| Bacterial pneumonia | 480.1 | rs147678978 | 3.71 | 3.80x10^-31^ |
| Bacterial pneumonia | 480.1 | rs138798329 | 3.70 | 9.83x10^-26^ |
| Bacterial pneumonia | 480.1 | rs145179853 | 3.45 | 4.68x10^-25^ |
| Bacterial pneumonia | 480.1 | rs35130797 | 2.17 | 4.83x10^-25^ |
| Bacterial pneumonia | 480.1 | rs1800503 | 2.17 | 5.66x10^-25^ |
| Bacterial pneumonia | 480.1 | rs34654194 | 2.16 | 7.19x10^-25^ |
| Bacterial pneumonia | 480.1 | rs34420782 | 2.16 | 7.48x10-^-25^ |
| Bacterial pneumonia | 480.1 | rs34268361 | 2.16 | 1.15x10^-25^ |
| Bacterial pneumonia | 480.1 | rs17139821 | 2.14 | 2.10x10^-24^ |
| Bacterial pneumonia | 480.1 | rs7802924 | 2.12 | 7.37x10^-24^ |
| Bacterial pneumonia | 480.1 | rs73480763 | 2.12 | 7.98x10^-24^ |
| Bacterial pneumonia | 480.1 | rs35003065 | 2.12 | 8.49x10^-24^ |
| Bacterial pneumonia | 480.1 | rs35820454 | 2.12 | 9.57x10^-24^ |
| Bacterial pneumonia | 480.1 | rs10237017 | 2.12 | 1.17x10^-23^ |
| Bacterial pneumonia | 480.1 | rs35715578 | 2.11 | 1.48x10^-23^ |
| Bacterial pneumonia | 480.1 | rs6943509 | 2.11 | 1.98x10^-23^ |
| Bacterial pneumonia | 480.1 | rs35987553 | 2.11 | 2.16x10^-23^ |
| Bacterial pneumonia | 480.1 | rs17544734 | 2.10 | 2.79x10^-23^ |
| Bacterial pneumonia | 480.1 | rs17545644 | 2.10 | 2.91x10^-23^ |
| Bacterial pneumonia | 480.1 | rs6961545 | 2.08 | 6.71x10^-23^ |
| Bacterial pneumonia | 480.1 | rs213972 | 2.08 | 7.84x10^-23^ |
| Bacterial pneumonia | 480.1 | rs10487364 | 2.08 | 8.29x10^-23^ |
| Bacterial pneumonia | 480.1 | rs79643487 | 2.07 | 2.23x10^-22^ |
| Bacterial pneumonia | 480.1 | rs1025342 | 2.06 | 2.85x10^-22^ |
| Bacterial pneumonia | 480.1 | rs17547485 | 2.04 | 1.32x10^-21^ |
| Bacterial pneumonia | 480.1 | rs17450482 | 2.04 | 1.40x10^-21^ |
| Bacterial pneumonia | 480.1 | rs213954 | 2.02 | 3.18x10^-21^ |
| Bacterial pneumonia | 480.1 | rs2518872 | 2.01 | 3.70x10^-21^ |
| Bacterial pneumonia | 480.1 | rs117998922 | 2.91 | 1.58x10^-19^ |
| Bacterial pneumonia | 480.1 | rs117606641 | 2.89 | 1.92x10^-18^ |
| Bacterial pneumonia | 480.1 | rs34638253 | 1.84 | 6.41x10^-18^ |
| Bacterial pneumonia | 480.1 | rs34159932 | 1.84 | 6.41x10^-18^ |
| Bacterial pneumonia | 480.1 | rs35141759 | 1.83 | 9.44x10^-18^ |
| Bacterial pneumonia | 480.1 | rs10487372 | 1.81 | 3.73x10^-17^ |
| Bacterial pneumonia | 480.1 | rs75359778 | 2.24 | 6.99x10^-16^ |
| Bacterial pneumonia | 480.1 | rs144286944 | 2.68 | 9.02x10^-16^ |
| Bacterial pneumonia | 480.1 | rs79584577 | 1.67 | 5.25x10^-15^ |
| Bacterial pneumonia | 480.1 | rs77532575 | 2.10 | 3.49x10^-14^ |
| Bacterial pneumonia | 480.1 | rs189240181 | 2.35 | 6.71x10^-13^ |
| Bacterial pneumonia | 480.1 | rs34363739 | 2.23 | 8.31x10^-11^ |
| Bacterial pneumonia | 480.1 | rs39308 | 1.44 | 2.43x10^-10^ |
| Bacterial pneumonia | 480.1 | rs39309 | 1.44 | 2.84x10^-10^ |
| Bacterial pneumonia | 480.1 | rs1989836 | 1.44 | 3.78x10^-10^ |
| Bacterial pneumonia | 480.1 | rs35453239 | 1.55 | 4.57x10^-10^ |
| Bacterial pneumonia | 480.1 | rs39310 | 1.43 | 7.02x10^-10^ |
| Bacterial pneumonia | 480.1 | rs76433139 | 2.75 | 7.26x10^-10^ |
| Bacterial pneumonia | 480.1 | rs39311 | 1.43 | 8.23x10^-10^ |
| Bacterial pneumonia | 480.1 | rs4730810 | 2.41 | 8.37x10^-10^ |
| Bacterial pneumonia | 480.1 | rs35596763 | 1.56 | 8.45x10^-10^ |
| Herpes simplex | 54 | rs148993397 | 4.44 | 9.57x10^-10^ |
| Staphylococcus infections | 41.1 | rs192146294 | 2.27 | 1.23x10^-9^ |
| Bacterial pneumonia | 480.1 | rs143583101 | 2.29 | 1.29x10^-9^ |
| Bacterial pneumonia | 480.1 | rs254538 | 1.41 | 1.90x10^-9^ |
| Bacterial pneumonia | 480.1 | rs6959515 | 1.41 | 1.94x10^-9^ |
| Mycoses | 117 | rs11779069 | 2.54 | 2.28x10^-9^ |
| Bacterial pneumonia | 480.1 | rs10487368 | 1.45 | 2.29x10^-9^ |
| Bacterial pneumonia | 480.1 | rs39314 | 1.41 | 2.51x10^-9^ |
| Bacterial pneumonia | 480.1 | rs2237724 | 1.45 | 2.53x10^-9^ |
| Bacterial pneumonia | 480.1 | rs39322 | 1.41 | 2.63x10^-9^ |
| Bacterial pneumonia | 480.1 | rs2188159 | 1.45 | 2.75x10^-9^ |
| Bacterial pneumonia | 480.1 | rs39321 | 1.41 | 2.83x10^-9^ |
| Bacterial pneumonia | 480.1 | rs213958 | 1.44 | 3.24x10^-9^ |
| Bacterial pneumonia | 480.1 | rs6466613 | 1.45 | 3.30x10^-9^ |
| Bacterial pneumonia | 480.1 | rs11770639 | 1.45 | 3.31x10^-9^ |
| Bacterial pneumonia | 480.1 | rs6969138 | 1.45 | 3.44x10^-9^ |
| Bacterial pneumonia | 480.1 | rs4148699 | 1.45 | 3.44x10^-9^ |
| Bacterial pneumonia | 480.1 | rs1557630 | 1.45 | 3.77x10^-9^ |
| Bacterial pneumonia | 480.1 | rs2027944 | 1.45 | 3.80x10^-9^ |
| Bacterial pneumonia | 480.1 | rs4148706 | 1.45 | 3.85x10^-9^ |
| Bacterial pneumonia | 480.1 | rs12529571 | 2.79 | 3.85x10^-9^ |
| Bacterial pneumonia | 480.1 | rs12538252 | 1.45 | 3.85x10^-9^ |
| Bacterial pneumonia | 480.1 | rs957461 | 1.45 | 3.99x10^-9^ |
| Bacterial pneumonia | 480.1 | rs1429566 | 1.45 | 3.99x10^-9^ |
| Bacterial pneumonia | 480.1 | rs39320 | 1.41 | 4.03x10^-9^ |
| Bacterial pneumonia | 480.1 | rs4148687 | 1.45 | 4.12x10^-9^ |
| Bacterial pneumonia | 480.1 | rs1896888 | 1.45 | 4.13x10^-9^ |
| Bacterial pneumonia | 480.1 | rs2027945 | 1.45 | 4.13x10^-9^ |
| HIV disease | 71 | rs4149909 | 3.42 | 4.40x10^-9^ |
| Bacterial pneumonia | 480.1 | rs4148686 | 1.44 | 4.48x10^-9^ |
| Bacterial pneumonia | 480.1 | rs10258791 | 1.41 | 4.49x10^-9^ |
| Mycoses | 117 | rs112971459 | 2.43 | 5.02x10^-9^ |
| Bacterial pneumonia | 480.1 | rs35388491 | 1.44 | 5.15x10^-9^ |
| Bacterial pneumonia | 480.1 | rs2237723 | 1.44 | 5.16x10^-9^ |
| Bacterial pneumonia | 480.1 | rs4148690 | 1.44 | 5.25x10^-9^ |
| Bacterial pneumonia | 480.1 | rs4148691 | 1.44 | 5.35x10^-9^ |
| Bacterial pneumonia | 480.1 | rs4148692 | 1.44 | 5.35x10^-9^ |
| Bacterial pneumonia | 480.1 | rs6951141 | 1.44 | 5.50x10^-9^ |
| Bacterial pneumonia | 480.1 | rs6957317 | 1.44 | 5.63x10^-9^ |
| Bacterial pneumonia | 480.1 | rs10242827 | 1.40 | 5.85x10^-9^ |
| Bacterial pneumonia | 480.1 | rs4148689 | 1.44 | 5.92x10^-9^ |
| Bacterial pneumonia | 480.1 | rs7793127 | 1.44 | 6.08x10^-9^ |
| Bacterial pneumonia | 480.1 | rs7779160 | 1.40 | 6.10x10^-9^ |
| Bacterial pneumonia | 480.1 | rs2402205 | 1.44 | 6.12x10^-9^ |
| Bacterial pneumonia | 480.1 | rs2402204 | 1.44 | 6.40x10^-9^ |
| Bacterial pneumonia | 480.1 | rs10230259 | 1.40 | 6.48x10^-9^ |
| Bacterial pneumonia | 480.1 | rs12673260 | 1.44 | 6.78x10^-9^ |
| Influenza | 481 | rs142537151 | 3.82 | 6.91x10^-9^ |
| Bacterial pneumonia | 480.1 | rs2283057 | 1.44 | 6.93x10^-9^ |
| Bacterial pneumonia | 480.1 | rs9886209 | 1.40 | 7.17x10^-9^ |
| Bacterial pneumonia | 480.1 | rs916728 | 1.40 | 7.18x10^-9^ |
| Bacterial pneumonia | 480.1 | rs6977764 | 1.44 | 7.21x10^-9^ |
| Bacterial pneumonia | 480.1 | rs10487369 | 1.44 | 7.21x10^-9^ |
| Bacterial pneumonia | 480.1 | rs4148696 | 1.44 | 7.27x10^-9^ |
| Mycoses | 117 | rs900052 | 2.40 | 7.30x10^-9^ |
| Bacterial pneumonia | 480.1 | rs916727 | 1.40 | 7.31x10^-9^ |
| Bacterial pneumonia | 480.1 | rs76644237 | 2.77 | 7.51x10^-9^ |
| Bacterial pneumonia | 480.1 | rs3808186 | 1.44 | 7.60x10^-9^ |
| Bacterial pneumonia | 480.1 | rs1896886 | 1.44 | 7.78x10^-9^ |
| Bacterial pneumonia | 480.1 | rs75912197 | 1.74 | 7.79x10^-9^ |
| Bacterial pneumonia | 480.1 | rs4148697 | 1.44 | 7.82x10^-9^ |
| Bacterial pneumonia | 480.1 | rs10255092 | 1.40 | 7.89x10^-9^ |
| Bacterial pneumonia | 480.1 | rs39325 | 1.40 | 7.98x10^-9^ |
| Meningitis | 320 | rs57665613 | 3.22 | 8.12x10^-9^ |
| Bacterial pneumonia | 480.1 | rs12533524 | 1.43 | 8.19x10^-9^ |
| Bacterial pneumonia | 480.1 | rs2283055 | 1.43 | 8.22x10^-9^ |
| Bacterial pneumonia | 480.1 | rs10953847 | 1.43 | 8.29x10^-9^ |
| Bacterial pneumonia | 480.1 | rs1896887 | 1.43 | 8.44x10^-9^ |
| Bacterial pneumonia | 480.1 | rs6963669 | 1.39 | 8.58x10^-9^ |
| Bacterial pneumonia | 480.1 | rs35608825 | 1.43 | 8.60x10^-9^ |
| Bacterial pneumonia | 480.1 | rs34683806 | 1.43 | 8.60x10^-9^ |
| Bacterial pneumonia | 480.1 | rs113121382 | 2.66 | 8.62x10^-9^ |
| Bacterial pneumonia | 480.1 | rs7786196 | 1.43 | 8.69x10^-9^ |
| Bacterial pneumonia | 480.1 | rs213956 | 1.43 | 8.75x10^-9^ |
| Meningitis | 320 | rs78071398 | 3.95 | 8.87x10^-9^ |
| Viral infection | 79 | rs77869329 | 2.48 | 8.96x10^-9^ |
| Viral infection | 79 | rs736962 | 2.48 | 8.96x10^-9^ |
| Viral infection | 79 | rs736961 | 2.48 | 8.96x10^-9^ |
| Viral infection | 79 | rs76728606 | 2.48 | 8.96x10^-9^ |
| Viral infection | 79 | rs77221291 | 2.48 | 8.96x10^-9^ |
| Bacterial pneumonia | 480.1 | rs2283056 | 1.43 | 8.97x10^-9^ |
| Bacterial pneumonia | 480.1 | rs4285411 | 1.39 | 8.97x10^-9^ |
| Bacterial pneumonia | 480.1 | rs4148688 | 1.43 | 9.03x10^-9^ |
| Bacterial pneumonia | 480.1 | rs34855237 | 1.43 | 9.16x10^-9^ |
| Bacterial pneumonia | 480.1 | rs2402228 | 1.43 | 9.40x10^-9^ |
| Mycoses | 117 | rs1277876 | 2.38 | 9.46x10^-9^ |
| Viral infection | 79 | rs2247438 | 2.45 | 9.66x10^-9^ |
| HIV disease | 71 | rs72755295 | 3.33 | 9.80x10^-9^ |
| Bacterial pneumonia | 480.1 | rs6466610 | 1.39 | 9.89x10^-9^ |
| Mycoses | 117 | rs2383921 | 2.38 | 1.00x10^-8^ |
| Meningitis | 320 | rs57818394 | 3.19 | 1.01x10^-8^ |
| Mycoses | 117 | rs1277875 | 2.38 | 1.02x10^-8^ |
| Meningitis | 320 | rs113123547 | 3.19 | 1.05x10^-8^ |
| Bacterial pneumonia | 480.1 | rs3757802 | 1.43 | 1.09x10^-8^ |
| Mycoses | 117 | rs139841939 | 2.37 | 1.10x10^-8^ |
| Meningitis | 320 | rs73403463 | 3.19 | 1.13x10^-8^ |
| Mycoses | 117 | rs2046181 | 2.37 | 1.15x10^-8^ |
| Mycoses | 117 | rs7018439 | 2.37 | 1.15x10^-8^ |
| Bacterial pneumonia | 480.1 | rs254537 | 1.39 | 1.16x10^-8^ |
| Mycoses | 117 | rs7823060 | 2.37 | 1.17x10^-8^ |
| Bacterial pneumonia | 480.1 | rs10264450 | 1.39 | 1.26x10^-8^ |
| Meningitis | 320 | rs10519083 | 3.17 | 1.26x10^-8^ |
| Bacterial pneumonia | 480.1 | rs10278953 | 1.39 | 1.37x10^-8^ |
| Viral infection | 79 | rs76357476 | 2.48 | 1.43x10^-8^ |
| Viral infection | 79 | rs36061079 | 2.48 | 1.44x10^-8^ |
| Viral infection | 79 | rs74713549 | 2.48 | 1.44x10^-8^ |
| Viral infection | 79 | rs79778361 | 2.48 | 1.44x10^-8^ |
| Herpes zoster | 53 | rs114514696 | 2.59 | 1.52x10^-8^ |
| Viral infection | 79 | rs2284670 | 2.47 | 1.53x10^-8^ |
| Bacterial pneumonia | 480.1 | rs116900901 | 1.73 | 1.63x10^-8^ |
| Bacterial pneumonia | 480.1 | rs9648982 | 1.39 | 1.65x10^-8^ |
| Influenza | 481 | rs146280264 | 3.57 | 1.67x10^-8^ |
| Bacterial pneumonia | 480.1 | rs149521175 | 1.72 | 1.69x10^-8^ |
| Herpes zoster | 53 | rs115500050 | 2.58 | 1.74x10^-8^ |
| Herpes zoster | 53 | rs79825409 | 2.58 | 1.78x10^-8^ |
| Bacterial pneumonia | 480.1 | rs213955 | 1.42 | 1.83x10^-8^ |
| Gram positive septicemia | 38.2 | rs2304535 | 3.88 | 1.84x10^-8^ |
| Herpes zoster | 53 | rs150554177 | 2.57 | 1.99x10^-8^ |
| Bacterial pneumonia | 480.1 | rs78905547 | 2.22 | 2.05x10^-8^ |
| Meningitis | 320 | rs77958030 | 3.16 | 2.25x10^-8^ |
| HIV disease | 71 | rs62470214 | 4.95 | 2.32x10^-8^ |
| Herpes simplex | 54 | rs5755132 | 2.19 | 2.34x10^-8^ |
| Gram positive septicemia | 38.2 | rs72960375 | 3.84 | 2.35x10^-8^ |
| Gram positive septicemia | 38.2 | rs72960366 | 3.84 | 2.35x10^-8^ |
| Gram positive septicemia | 38.2 | rs17022172 | 3.84 | 2.36x10^-8^ |
| Herpes zoster | 53 | rs139206024 | 3.73 | 2.37x10^-8^ |
| Gram positive septicemia | 38.2 | rs2043703 | 3.83 | 2.44x10^-8^ |
| Gram positive septicemia | 38.2 | rs72943804 | 3.83 | 2.44x10^-8^ |
| Gram positive septicemia | 38.2 | rs72943833 | 3.83 | 2.44x10^-8^ |
| Gram positive septicemia | 38.2 | rs80127877 | 3.83 | 2.44x10^-8^ |
| Meningitis | 320 | rs72742268 | 4.26 | 2.48x10^-8^ |
| Herpes simplex | 54 | rs5749894 | 2.19 | 2.51x10^-8^ |
| Herpes simplex | 54 | rs4605473 | 2.19 | 2.51x10^-8^ |
| Other infectious and parasitic diseases | 136 | rs116274786 | 4.96 | 2.53x10^-8^ |
| Viral infection | 79 | rs12413743 | 2.43 | 2.63x10^-8^ |
| Bacterial pneumonia | 480.1 | rs7786419 | 1.43 | 2.67x10^-8^ |
| Viral infection | 79 | rs12416424 | 2.43 | 2.75x10^-8^ |
| Bacterial pneumonia | 480.1 | rs10227378 | 1.38 | 2.80x10^-8^ |
| Meningitis | 320 | rs78009408 | 3.14 | 2.80x10^-8^ |
| Gram positive septicemia | 38.2 | rs17022100 | 3.76 | 2.83x10^-8^ |
| Dermatophytosis / Dermatomycosis | 110 | rs41302744 | 2.20 | 2.86x10^-8^ |
| Meningitis | 320 | rs76465403 | 3.14 | 2.87x10^-8^ |
| Meningitis | 320 | rs75569052 | 3.13 | 2.93x10^-8^ |
| Meningitis | 320 | rs4843893 | 3.65 | 3.14x10^-8^ |
| Meningitis | 320 | rs55668004 | 3.13 | 3.17x10^-8^ |
| Other CNS infection and poliomyelitis | 324 | rs61950211 | 3.67 | 3.21x10^-8^ |
| Bacterial pneumonia | 480.1 | rs12536356 | 1.38 | 3.27x10^-8^ |
| Meningitis | 320 | rs55676216 | 3.13 | 3.28x10^-8^ |
| Meningitis | 320 | rs74014424 | 3.13 | 3.31x10^-8^ |
| Meningitis | 320 | rs17322852 | 3.13 | 3.31x10^-8^ |
| Meningitis | 320 | rs74014423 | 3.12 | 3.43x10^-8^ |
| Meningitis | 320 | rs74014432 | 3.12 | 3.45x10^-8^ |
| Meningitis | 320 | rs58735171 | 3.12 | 3.60x10^-8^ |
| Meningitis | 320 | rs115951300 | 3.12 | 3.63x10^-8^ |
| Meningitis | 320 | rs77618710 | 3.11 | 3.64x10^-8^ |
| Meningitis | 320 | rs79782076 | 3.11 | 3.65x10^-8^ |
| Meningitis | 320 | rs58892007 | 3.10 | 3.70x10^-8^ |
| Meningitis | 320 | rs56078881 | 3.11 | 3.71x10^-8^ |
| Viral infection | 79 | rs77208736 | 2.43 | 3.71x10^-8^ |
| Meningitis | 320 | rs74014429 | 3.11 | 3.75x10^-8^ |
| Meningitis | 320 | rs56165109 | 3.11 | 3.75x10^-8^ |
| Meningitis | 320 | rs2178098 | 3.11 | 3.75x10^-8^ |
| Meningitis | 320 | rs73403471 | 3.09 | 3.78x10^-8^ |
| Herpes zoster | 53 | rs117652069 | 3.05 | 3.79x10^-8^ |
| Meningitis | 320 | rs1918665 | 3.10 | 3.82x10^-8^ |
| Meningitis | 320 | rs73403474 | 3.08 | 3.90x10^-8^ |
| Viral infection | 79 | rs2268352 | 2.43 | 3.95x10^-8^ |
| Meningitis | 320 | rs16958667 | 3.11 | 3.99x10^-8^ |
| Meningitis | 320 | rs74014435 | 3.11 | 3.99x10^-8^ |
| Meningitis | 320 | rs74014437 | 3.11 | 3.99x10^-8^ |
| Viral infection | 79 | rs2901309 | 2.40 | 4.10x10^-8^ |
| Meningitis | 320 | rs75876689 | 3.10 | 4.15x10^-8^ |
| Other CNS infection and poliomyelitis | 324 | rs13296786 | 3.95 | 4.16x10^-8^ |
| Bacterial pneumonia | 480.1 | rs12706151 | 1.37 | 4.18x10^-8^ |
| Viral infection | 79 | rs61818960 | 2.73 | 4.18x10^-8^ |
| Viral infection | 79 | rs117966582 | 2.42 | 4.22x10^-8^ |
| Viral infection | 79 | rs17103661 | 2.42 | 4.22x10^-8^ |
| Gram positive septicemia | 38.2 | rs76188060 | 3.69 | 4.22x10^-8^ |
| Meningitis | 320 | rs74014431 | 3.10 | 4.22x10^-8^ |
| Viral infection | 79 | rs79709304 | 2.42 | 4.34x10^-8^ |
| Other CNS infection and poliomyelitis | 324 | rs140636131 | 4.38 | 4.35x10^-8^ |
| Herpes zoster | 53 | rs117755556 | 2.43 | 4.36x10^-8^ |
| Meningitis | 320 | rs16958673 | 3.10 | 4.45x10^-8^ |
| Viral infection | 79 | rs77571505 | 2.42 | 4.49x10^-8^ |
| Viral infection | 79 | rs77637308 | 2.42 | 4.53x10^-8^ |
| Viral infection | 79 | rs117998404 | 2.42 | 4.60x10^-8^ |
| Viral infection | 79 | rs77872346 | 2.42 | 4.61x10^-8^ |
| Viral infection | 79 | rs4133111 | 2.42 | 4.64x10^-8^ |
| Viral infection | 79 | rs117396934 | 2.42 | 4.64x10^-8^ |
| Viral infection | 79 | rs4514345 | 2.41 | 4.67x10^-8^ |
| Viral infection | 79 | rs79999535 | 2.41 | 4.71x10^-8^ |
| Viral infection | 79 | rs76912423 | 2.41 | 4.74x10^-8^ |
| Viral infection | 79 | rs77927946 | 2.41 | 4.76x10^-8^ |
| Viral infection | 79 | rs75759547 | 2.41 | 4.76x10^-8^ |
| Viral infection | 79 | rs77022052 | 2.41 | 4.76x10^-8^ |
| Viral infection | 79 | rs76051814 | 2.41 | 4.78x10^-8^ |
| Viral infection | 79 | rs28549398 | 2.41 | 4.78x10^-8^ |
| Viral infection | 79 | rs79412091 | 2.41 | 4.78x10^-8^ |
| Viral infection | 79 | rs28616878 | 2.41 | 4.78x10^-8^ |
| Viral infection | 79 | rs76453308 | 2.41 | 4.78x10^-8^ |
| Viral infection | 79 | rs74661887 | 2.41 | 4.78x10^-8^ |
| Viral infection | 79 | rs74436549 | 2.41 | 4.78x10^-8^ |
| Viral infection | 79 | rs80288452 | 2.41 | 4.78x10^-8^ |
| Viral infection | 79 | rs80350367 | 2.41 | 4.78x10^-8^ |
| Dermatophytosis / Dermatomycosis | 110 | rs10207907 | 1.79 | 4.80x10^-8^ |
| Viral infection | 79 | rs79902378 | 2.41 | 4.87x10^-8^ |
| Viral infection | 79 | rs78271758 | 2.41 | 4.87x10^-8^ |
| Viral infection | 79 | rs75992056 | 2.41 | 4.87x10^-8^ |
| Viral infection | 79 | rs79127377 | 2.41 | 4.87x10^-8^ |
| Herpes simplex | 54 | rs143539215 | 3.00 | 4.87x10^-8^ |
| Viral infection | 79 | rs78036758 | 2.41 | 4.95x10^-8^ |
| Viral infection | 79 | rs76970830 | 2.41 | 4.95x10^-8^ |
| Viral infection | 79 | rs75796417 | 2.41 | 4.95x10^-8^ |
| Meningitis | 320 | rs73403462 | 3.07 | 4.96x10^-8^ |

**Table S2.** Proteins that are post-translationally modified by phosphorylation among 70 experiment-wide or individual ID trait significance.

**Protein**

AGTR1, AKIRIN2, ALX4, ANK2, ASPSCR1, ATG9A, AVEN, AVIL, CDK5, CERS3, CHMP5, CRYL1, DNAJC17, EIF3M, FABP4, FAM208A, GPRIN1, HIP1, LIMK2, MAATS1, MAPK8IP2, MYO1C, NDUFA4, NELFB, NSUN5, NUDT5, PIP5K1A, PSMG1, PTPN4, RAD18, RAD50, SLC35F6, STAP2, TBK1, TOR4A, TXLNB, TXNL1, USP44, VWA5B1, WIPF1

**Table S3.** Proteins that are post-translationally modified by acetylation among 70 experiment-wide or individual ID trait significance.

**Protein**

ASPSCR1, ATG9A, CDK5, EIF3M, FABP4, FAM208A, MYO1C, NDUFA4, NELFB, NSUN5, NUDT5, PSMG1, PTCD3, RAD18, RAD50, TNNC2

**Table S4.** ID phenotypes nominally-associated (p < 0.05) with significant gene-level associations for individual ID traits, cases > 100.

| **Gene** | **PheCode** | **Phenotype** | **Cases** | **Controls** | **Ancestry** | **Odds ratio** | **P value** |
| --- | --- | --- | --- | --- | --- | --- | --- |
| IKZF5 | 079 | Viral infection | 1,811 | 20,904 | European | 0.93 | 2.8x10^-3^ |
| IKZF5 | 038.1 | Gram negative septicemia | 820 | 19,844 | European | 0.91 | 1.11x10^-2^ |
| IKZF5 | 481 | Influenza | 272 | 18,054 | European | 0.86 | 1.98x10^-2^ |
| IKZF5 | 112 | Candidiasis | 2,284 | 21,426 | European | 0.95 | 3.17x10^-2^ |
| IKZF5 | 008 | Intestinal infection | 1,608 | 24,187 | European | 0.95 | 4.32x10^-2^ |
| IKZF5 | 041.9 | Infection with drug-resistant microorganisms | 893 | 19,844 | European | 0.93 | 4.65x10^-2^ |
| AKIRIN2 | 041.9 | Infection with drug-resistant microorganisms | 893 | 19,844 | European | 0.90 | 2.20x10^-3^ |
| AKIRIN2 | 041.1 | *Staphylococcus* infections | 2,180 | 19,844 | European | 0.94 | 1.08x10^-2^ |
| AKIRIN2 | 480.1 | Bacterial pneumonia | 862 | 18,054 | European | 0.93 | 3.27x10^-2^ |
| AKIRIN2 | 041.8 | *Helicobacter pylori* infection | 150 | 19,844 | European | 1.21 | 3.86x10^-2^ |
| PSMG1 | 038.2 | Gram positive septicemia | 613 | 19,844 | European | 0.84 | 2.16x10^-5^ |
| PSMG1 | 041.4 | *Escherichia coli* infection | 1,231 | 19,844 | European | 0.92 | 5.42x10^-3^ |
| PSMG1 | 136 | Other infectious and parasitic diseases | 746 | 24,770 | European | 0.93 | 4.75x10^-2^ |
| AGTR1 | 070.3 | Viral hepatitis C | 808 | 20,904 | European | 1.10 | 7.54x10^-3^ |
| AGTR1 | 110 | Dermatophytosis and dermatomycosis | 2,192 | 21,426 | European | 1.05 | 1.90x10^-2^ |
| AGTR1 | 010 | Tuberculosis | 156 | 19,844 | European | 0.81 | 2.33x10^-2^ |
| SLC35F6 | 041.9 | Infection with drug-resistant microorganisms | 893 | 19,844 | European | 0.87 | 1.57x10^-4^ |
| SLC35F6 | 054 | Herpes simplex | 610 | 20,904 | European | 0.88 | 2.62x10^-3^ |
| SLC35F6 | 053 | Herpes zoster | 898 | 20,904 | European | 0.91 | 9.00x10^-3^ |
| SLC35F6 | 117 | Mycoses | 627 | 21,426 | European | 0.90 | 1.66x10^-2^ |
| SLC35F6 | 324 | Other CNS infection and poliomyelitis | 136 | 25,170 | European | 1.20 | 2.43x10^-2^ |
| NDUFAA4 | 070.2 | Viral hepatitis B | 166 | 20,904 | European | 1.28 | 2.07x10^-4^ |
| NDUFAA4 | 110 | Dermatophytosis and dermatomycosis | 2,192 | 21,426 | European | 1.07 | 2.08x10^-3^ |
| NDUFAA4 | 136 | Other infectious and parasitic diseases | 746 | 24,770 | European | 1.10 | 8.84x10^-3^ |
| NDUFAA4 | 078 | Viral warts and HPV | 1,152 | 20,904 | European | 1.08 | 1.32x10^-2^ |
| NDUFAA4 | 112 | Candidiasis | 2,284 | 21,426 | European | 1.05 | 1.90x10^-2^ |
| NDUFAA4 | 010 | Tuberculosis | 156 | 19,844 | European | 0.82 | 2.28x10^-2^ |
| NDUFAA4 | 038.1 | Gram negative septicemia | 820 | 19,844 | European | 0.92 | 2.90x10^-2^ |
| NDUFAA4 | 053 | Herpes zoster | 898 | 20,904 | European | 1.07 | 3.50x10^-2^ |
| C10orf120 | 117 | Mycoses | 627 | 21,426 | European | 1.14 | 1.49x10^-4^ |
| C10orf120 | 079 | Viral infection | 1,811 | 20,904 | European | 1.09 | 3.60x10^-4^ |
| C10orf120 | 038.2 | Gram positive septicemia | 613 | 19,844 | European | 1.13 | 5.32x10^-4^ |
| C10orf120 | 480.1 | Bacterial pneumonia | 862 | 18,054 | European | 1.10 | 2.48x10^-3^ |
| C10orf120 | 054 | Herpes simplex | 610 | 20,904 | European | 1.11 | 3.70x10^-3^ |
| C10orf120 | 041.4 | *Escherichia coli* infection | 1,231 | 19,844 | European | 1.08 | 7.88x10^-3^ |
| C10orf120 | 041.2 | *Streptococcus* infection | 1,262 | 19,844 | European | 1.07 | 2.33x10^-3^ |
| C10orf120 | 041.9 | Infection with drug-resistant microorganisms | 893 | 19,844 | European | 1.07 | 4.23x10^-3^ |
| C10orf120 | 053 | Herpes zoster | 898 | 20,904 | European | 1.07 | 4.65x10^-3^ |
| RAD18 | 070.2 | Viral hepatitis B | 166 | 20,904 | European | 1.28 | 3.62x10^-3^ |
| RAD18 | 041.1 | *Staphylococcus* infections | 2,180 | 19,844 | European | 1.06 | 1.53x10^-2^ |
| RAD18 | 070.3 | Viral hepatitis C | 808 | 20,904 | European | 1.09 | 2.04x10^-2^ |
| RAD18 | 071 | Human immunodeficiency virus (HIV) | 196 | 20,904 | European | 0.86 | 2.62x10^-2^ |
| MAPK8IP2 | 038.1 | Gram negative septicemia | 820 | 19,844 | European | 1.10 | 8.17x10^-3^ |
| AVIL | 070.2 | Viral hepatitis B | 166 | 20,904 | European | 1.20 | 2.03x10^-2^ |
| AVIL | 053 | Herpes zoster | 898 | 20,904 | European | 0.93 | 4.18x10^-2^ |
| STAP2 | 008 | Intestinal infection | 1,608 | 24,187 | European | 1.06 | 2.46x10^-2^ |
| STAP2 | 053 | Herpes zoster | 898 | 20,904 | European | 1.08 | 3.00x10^-2^ |
| STAP2 | 481 | Influenza | 272 | 18,054 | European | 1.13 | 3.55x10^-2^ |
| STAP2 | 320 | Meningitis | 114 | 25,170 | European | 0.83 | 3.83x10^-2^ |
| TXLNB | 054 | Herpes simplex | 610 | 20,904 | European | 0.91 | 2.20x10^-2^ |
| TXLNB | 112 | Candidiasis | 2,284 | 21,426 | European | 1.05 | 4.10x10^-2^ |
| CLDN20 | 008 | Intestinal infection | 1,608 | 24,187 | European | 0.94 | 1.92x10^-2^ |
| IGF2 | 041.1 | *Staphylococcus* infections | 2,180 | 19,844 | European | 0.94 | 9.50x10^-3^ |
| IGF2 | 110 | Dermatophytosis and dermatomycosis | 2,192 | 21,426 | European | 1.06 | 1.21x10^-2^ |
| IGF2 | 010 | Tuberculosis | 156 | 19,844 | European | 0.82 | 1.37x10^-2^ |
| IGF2 | 320 | Meningitis | 114 | 25,170 | European | 1.18 | 4.40x10^-2^ |
| CERS3 | 041.1 | *Staphylococcus* infections | 2,180 | 19,844 | European | 1.07 | 2.74x10^-3^ |
| CERS3 | 008 | Intestinal infection | 1,608 | 24,187 | European | 1.08 | 3.52x10^-3^ |
| CERS3 | 480.1 | Bacterial pneumonia | 862 | 18,054 | European | 1.09 | 1.06x10^-2^ |
| CERS3 | 481 | Influenza | 272 | 18,054 | European | 1.14 | 2.84x10^-2^ |
| CERS3 | 071 | Human immunodeficiency virus (HIV) | 196 | 20,904 | European | 0.86 | 3.58x10^-2^ |
| CERS3 | 038.2 | Gram positive septicemia | 613 | 19,844 | European | 1.09 | 3.89x10^-2^ |
| TOR4A | 112 | Candidiasis | 2,284 | 21,426 | European | 0.93 | 5.21x10^-4^ |
| TOR4A | 041.1 | *Staphylococcus* infections | 2,180 | 19,844 | European | 0.93 | 1.43x10^-3^ |
| TOR4A | 038.1 | Gram negative septicemia | 820 | 19,844 | European | 0.93 | 2.86x10^-2^ |
| TOR4A | 041.9 | Infection with drug-resistant microorganisms | 893 | 19,844 | European | 0.93 | 3.02x10^-2^ |
| TOR4A | 038.2 | Gram positive septicemia | 613 | 19,844 | European | 0.92 | 3.91x10^-2^ |
| FAM166A | 112 | Candidiasis | 2,284 | 21,426 | European | 1.07 | 8.28x10^-4^ |
| FAM166A | 041.1 | *Staphylococcus* infections | 2,180 | 19,844 | European | 1.07 | 2.43x10^-3^ |
| FAM166A | 038.1 | Gram negative septicemia | 820 | 19,844 | European | 1.09 | 9.95x10^-3^ |
| C9orf173 | 112 | Candidiasis | 2,284 | 21,426 | European | 1.07 | 2.10x10^-3^ |
| C9orf173 | 041.1 | *Staphylococcus* infections | 2,180 | 19,844 | European | 1.07 | 4.07x10^-3^ |
| C9orf173 | 038.1 | Gram negative septicemia | 820 | 19,844 | European | 1.09 | 1.18x10^-2^ |
| C9orf173 | 041.9 | Infection with drug-resistant microorganisms | 893 | 19,844 | European | 1.07 | 4.20x10^-2^ |
| PIP5K1A | 041.1 | *Staphylococcus* infections | 2,180 | 19,844 | European | 1.06 | 9.99x10^-3^ |
| NELFB | 041.1 | *Staphylococcus* infections | 2,180 | 19,844 | European | 0.93 | 1.34x10^-3^ |
| NELFB | 112 | Candidiasis | 2,284 | 21,426 | European | 0.94 | 4.24x10^-3^ |
| NELFB | 041.9 | Infection with drug-resistant microorganisms | 893 | 19,844 | European | 0.92 | 1.81x10^-2^ |
| NELFB | 038.1 | Gram negative septicemia | 820 | 19,844 | European | 0.93 | 3.10x10^-2^ |
| NELFB | 038.2 | Gram positive septicemia | 613 | 19,844 | European | 0.92 | 4.86x10^-2^ |
| AVEN | 041.2 | *Streptococcus* infection | 1,262 | 19,844 | European | 0.92 | 3.94x10^-3^ |
| AVEN | 112 | Candidiasis | 2,284 | 21,426 | European | 0.94 | 7.68x10^-3^ |
| AVEN | 038.2 | Gram positive septicemia | 613 | 19,844 | European | 0.91 | 1.41x10^-2^ |
| AVEN | 041.1 | *Staphylococcus* infections | 2,180 | 19,844 | European | 0.95 | 2.65x10^-2^ |
| TM7SF3 | 041.4 | *Escherichia coli* infection | 1,231 | 19,844 | European | 1.07 | 1.47x10^-2^ |
| TM7SF3 | 071 | Human immunodeficiency virus (HIV) | 196 | 20,904 | European | 1.17 | 1.62x10^-2^ |
| TM7SF3 | 041.2 | *Streptococcus* infection | 1,262 | 19,844 | European | 1.06 | 2.75x10^-2^ |
| RAD50 | 053 | Herpes zoster | 898 | 20,904 | European | 0.90 | 4.63x10^-3^ |
| RAD50 | 041.9 | Infection with drug-resistant microorganisms | 893 | 19,844 | European | 1.08 | 2.69x10^-2^ |
| RAD50 | 041.2 | *Streptococcus* infection | 1,262 | 19,844 | European | 1.06 | 2.94x10^-2^ |
| ZNF577 | 010 | Tuberculosis | 156 | 19,844 | European | 0.82 | 8.26x10^-3^ |
| ZNF577 | 117 | Mycoses | 627 | 21,426 | European | 1.11 | 1.21x10^-2^ |
| ZNF577 | 112 | Candidiasis | 2,284 | 21,426 | European | 1.06 | 1.62x10^-2^ |
| ZNF577 | 320 | Meningitis | 114 | 25,170 | European | 1.23 | 2.20x10^-2^ |
| ZNF577 | 324 | Other CNS infection and poliomyelitis | 136 | 25,170 | European | 1.21 | 4.33x10^-2^ |
| ZNF649 | 038.1 | Gram negative septicemia | 820 | 19,844 | European | 0.89 | 6.65x10^-4^ |
| ZNF649 | 010 | Tuberculosis | 156 | 19,844 | European | 0.78 | 9.24x10^-4^ |
| ZNF649 | 324 | Other CNS infection and poliomyelitis | 136 | 25,170 | European | 1.29 | 6.41x10^-3^ |
| ZNF649 | 320 | Meningitis | 114 | 25,170 | European | 1.33 | 1.87x10^-2^ |
| ZNF649 | 041.2 | *Streptococcus* infection | 1,262 | 19,844 | European | 0.94 | 2.11x10^-2^ |
| ZNF649 | 071 | Human immunodeficiency virus (HIV) | 196 | 20,904 | European | 0.86 | 2.32x10^-2^ |
| SETD9 | 041.4 | *Escherichia coli* infection | 1,231 | 19,844 | European | 1.08 | 1.80x10^-2^ |
| SETD9 | 041.2 | *Streptococcus* infection | 1,262 | 19,844 | European | 1.07 | 1.87x10^-2^ |
| AC022431.1 | 041.2 | *Streptococcus* infection | 1,262 | 19,844 | European | 0.91 | 2.25x10^-3^ |
| AC022431.1 | 008 | Intestinal infection | 1,608 | 24,187 | European | 1.07 | 1.09x10^-2^ |
| AC022431.1 | 038.2 | Gram positive septicemia | 613 | 19,844 | European | 0.90 | 2.14x10^-2^ |
| AC022431.1 | 010 | Tuberculosis | 156 | 19,844 | European | 1.16 | 4.47x10^-2^ |
| MYO1C | 041.9 | Infection with drug-resistant microorganisms | 893 | 19,844 | European | 0.86 | 1.54x10^-3^ |
| NUDT5 | 070.2 | Viral hepatitis B | 166 | 20,904 | European | 0.74 | 2.27x10^-4^ |
| NUDT5 | 071 | Human immunodeficiency virus (HIV) | 196 | 20,904 | European | 0.78 | 8.30x10^-4^ |
| NUDT5 | 481 | Influenza | 272 | 18,054 | European | 1.19 | 4.99x10^-3^ |
| NUDT5 | 041.1 | *Staphylococcus* infections | 2,180 | 19,844 | European | 0.94 | 8.30x10^-3^ |
| NUDT5 | 117 | Mycoses | 627 | 21,426 | European | 0.91 | 2.33x10^-2^ |
| NUDT5 | 041.8 | *Helicobacter pylori* infection | 150 | 19,844 | European | 1.19 | 3.38x10^-2^ |
| NUDT5 | 041.9 | Infection with drug-resistant microorganisms | 893 | 19,844 | European | 0.93 | 4.74x10^-2^ |
| MAATS1 | 480.1 | Bacterial pneumonia | 112 | 2,991 | African | 0.76 | 1.96x10^-2^ |
| MAATS1 | 112 | Candidiasis | 563 | 3,330 | African | 0.89 | 2.03x10^-2^ |
| MAATS1 | 131 | Protozoan infection | 158 | 4,054 | African | 0.82 | 3.32x10^-2^ |
| PTPN4 | 041.1 | *Staphylococcus* infections | 2,180 | 19,844 | European | 0.93 | 1.27x10^-3^ |
| PTPN4 | 112 | Candidiasis | 2,284 | 21,426 | European | 0.94 | 5.91x10^-3^ |
| PTPN4 | 041.9 | Infection with drug-resistant microorganisms | 893 | 19,844 | European | 0.92 | 1.44x10^-2^ |
| WIPF1 | 480.1 | Bacterial pneumonia | 862 | 18,054 | European | 0.90 | 3.36x10^-3^ |
| WIPF1 | 070.2 | Viral hepatitis B | 166 | 20,904 | European | 0.83 | 2.25x10^-2^ |
| ALX4 | 010 | Tuberculosis | 156 | 19,844 | European | 0.76 | 1.05x10^-3^ |
| ALX4 | 110 | Dermatophytosis and dermatomycosis | 2,192 | 21,426 | European | 1.06 | 5.32x10^-3^ |
| ALX4 | 041.1 | *Staphylococcus* infections | 2,180 | 19,844 | European | 1.06 | 1.41x10^-2^ |
| ALX4 | 041.8 | *Helicobacter pylori* infection | 150 | 19,844 | European | 1.18 | 4.24x10^-2^ |
| ALX4 | 041.9 | Infection with drug-resistant microorganisms | 893 | 19,844 | European | 1.07 | 2.15x10^-2^ |
| C22orf31 | 481 | Influenza | 272 | 18,054 | European | 0.82 | 2.01x10^-3^ |
| C22orf31 | 136 | Other infectious and parasitic diseases | 746 | 24,770 | European | 0.90 | 6.13x10^-3^ |
| C22orf31 | 041.9 | Infection with drug-resistant microorganisms | 893 | 19,844 | European | 0.92 | 2.01x10^-2^ |
| C22orf31 | 070.2 | Viral hepatitis B | 166 | 20,904 | European | 0.84 | 2.81x10^-2^ |
| C22orf31 | 079 | Viral infection | 1,811 | 20,904 | European | 0.95 | 3.01x10^-2^ |
| C22orf31 | 041.2 | *Streptococcus* infection | 1,262 | 19,844 | European | 0.94 | 3.09x10^-2^ |
| IL2RA | 041.1 | *Staphylococcus* infections | 2,180 | 19,844 | European | 1.07 | 3.09x10^-3^ |
| IL2RA | 480.1 | Bacterial pneumonia | 862 | 18,054 | European | 1.10 | 4.80x10^-2^ |
| IL2RA | 481 | Influenza | 272 | 18,054 | European | 0.87 | 2.77x10^-2^ |
| IL2RA | 041.4 | *Escherichia coli* infection | 1,231 | 19,844 | European | 1.06 | 3.69x10^-2^ |
| VWA5B1 | 071 | Human immunodeficiency virus (HIV) | 139 | 3,241 | African | 0.84 | 4.08x10^-2^ |
| ATP6V1C2 | 041.9 | Infection with drug-resistant microorganisms | 165 | 3,337 | African | 1.22 | 2.17x10^-2^ |
| ATP6V1C2 | 112 | Candidiasis | 563 | 3,330 | African | 1.10 | 4.02x10^-2^ |
| FAM20A | 117 | Mycoses | 627 | 21,426 | European | 0.89 | 4.22x10^-3^ |
| FAM20A | 008 | Intestinal infection | 1,608 | 24,187 | European | 0.93 | 6.72x10^-3^ |
| ASPSCR1 | 038.2 | Gram positive septicemia | 613 | 19,844 | European | 0.87 | 4.53x10^-4^ |
| ASPSCR1 | 079 | Viral infection | 1,811 | 20,904 | European | 1.07 | 6.20x10^-3^ |
| ASPSCR1 | 054 | Herpes simplex | 610 | 20,904 | European | 1.10 | 1.97x10^-2^ |
| ASPSCR1 | 320 | Meningitis | 114 | 25,170 | European | 0.85 | 3.72x10^-2^ |
| FAM208A | 041.1 | *Staphyloccocus* infections | 358 | 3,337 | African | 1.17 | 6.03x10^-3^ |
| FAM208A | 038.1 | Gram negative septicemia | 139 | 3,337 | African | 1.27 | 8.45x10^-3^ |
| FAM208A | 038.2 | Gram positive septicemia | 114 | 3,337 | African | 1.29 | 1.10x10^-2^ |
| TBK1 | 041.9 | Infection with drug-resistant microorganisms | 165 | 3,337 | African | 1.32 | 1.83x10^-3^ |
| TBK1 | 136 | Other infectious and parasitic diseases | 173 | 4,054 | African | 1.29 | 1.99x10^-3^ |
| TBK1 | 038.2 | Gram positive septicemia | 114 | 3,337 | African | 1.31 | 9.81x10^-3^ |
| TBK1 | 053 | Herpes zoster | 141 | 3,241 | African | 1.26 | 1.03x10^-2^ |
| TBK1 | 038.1 | Gram negative septicemia | 139 | 3,337 | African | 1.26 | 1.27x10^-2^ |
| TBK1 | 110 | Dermatophytosis and dermatomycosis | 654 | 3,330 | African | 1.11 | 2.10x10^-2^ |
| TBK1 | 041.1 | *Staphyloccocus* infections | 358 | 3,337 | African | 1.14 | 2.45x10^-2^ |
| TBK1 | 480.1 | Bacterial pneumonia | 112 | 2,991 | African | 1.26 | 2.71x10^-2^ |
| DNAJC5G | 136 | Other infectious and parasitic diseases | 746 | 24,770 | European | 1.06 | 2.83x10^-5^ |
| DNAJC5G | 041.9 | Infection with drug-resistant microorganisms | 893 | 19,844 | European | 1.06 | 3.513x10^-5^ |
| DNAJC5G | 078 | Viral warts and HPV | 1,152 | 20,904 | European | 1304 | 5.39x10^-3^ |
| DNAJC5G | 041.1 | *Staphylococcus* infections | 2,180 | 19,844 | European | 1.04 | 1.09x10^-2^ |
| DNAJC5G | 070.2 | Viral hepatitis B | 166 | 20,904 | European | 1.06 | 2.22x10^-2^ |
| DNAJC5G | 038.2 | Gram positive septicemia | 613 | 19,844 | European | 1.04 | 2.39x10^-2^ |
| DNAJC5G | 112 | Candidiasis | 2,284 | 21,426 | European | 1.03 | 2.44x10^-2^ |
| FABP4 | 481 | Influenza | 272 | 18,054 | European | 0.83 | 1.08x10^-2^ |
| FABP4 | 071 | Human immunodeficiency virus (HIV) | 196 | 20,904 | European | 0.94 | 1.23x10^-2^ |
| FABP4 | 079 | Viral infection | 1,811 | 20,904 | European | 0.94 | 1.63x10^-2^ |
| ANK2 | 038.1 | Gram negative septicemia | 820 | 19,844 | European | 1.13 | 2.56x10^-3^ |
| ANK2 | 041.8 | *Helicobacter pylori* infection | 150 | 19,844 | European | 0.86 | 3.59x10^-2^ |
| ANK2 | 112 | Candidiasis | 2,284 | 21,426 | European | 1.05 | 3.87x10^-2^ |
| ANK2 | 070.3 | Viral hepatitis C | 808 | 20,904 | European | 0.93 | 4.41x10^-2^ |
| HIP1 | 041.4 | *Escherichia coli* infection | 243 | 3,337 | African | 1.27 | 2.49x10^-4^ |
| HIP1 | 136 | Other infectious and parasitic diseases | 173 | 4,054 | African | 1.26 | 1.49x10^-3^ |
| HIP1 | 041.1 | *Staphyloccocus* infections | 358 | 3,337 | African | 1.19 | 1.63x10^-3^ |
| HIP1 | 480.1 | Bacterial pneumonia | 112 | 2,991 | African | 1.31 | 3.90x10^-3^ |
| HIP1 | 038.2 | Gram positive septicemia | 114 | 3,337 | African | 1.30 | 4.40x10^-3^ |
| C11orf53 | 110 | Dermatophytosis and dermatomycosis | 654 | 3,330 | African | 0.86 | 1.94x10^-4^ |
| C11orf53 | 112 | Candidiasis | 563 | 3,330 | African | 0.88 | 4.74x10^-3^ |
| C11orf53 | 041.4 | *Escherichia coli* infection | 243 | 3,337 | African | 0.84 | 5.95x10^-3^ |
| C11orf53 | 070.3 | Viral hepatitis C | 150 | 3,241 | African | 1.23 | 3.44x10^-2^ |
| C11orf53 | 008 | Intestinal infection | 368 | 4,060 | African | 0.90 | 3.48x10^-2^ |
| C11orf53 | 480.1 | Bacterial pneumonia | 112 | 2,991 | African | 0.82 | 3.52x10^-2^ |
| C11orf53 | 131 | Protozoan infection | 158 | 4,054 | African | 0.86 | 4.46x10^-2^ |
| EPCAM | 112 | Candidiasis | 2,284 | 21,426 | European | 1.07 | 2.92x10^-3^ |
| EPCAM | 054 | Herpes simplex | 610 | 20,904 | European | 1.11 | 3.62x10^-3^ |
| EPCAM | 038.1 | Gram negative septicemia | 820 | 19,844 | European | 1.09 | 1.01x10^-2^ |
| EPCAM | 038.2 | Gram positive septicemia | 613 | 19,844 | European | 1.09 | 3.26x10^-2^ |
| EPCAM | 041.9 | Infection with drug-resistant microorganisms | 893 | 19,844 | European | 1.07 | 3.36x10^-2^ |
| EPCAM | 136 | Other infectious and parasitic diseases | 746 | 24,770 | European | 1.07 | 4.26x10^-2^ |
| PROX2 | 054 | Herpes simplex | 610 | 20,904 | European | 0.87 | 1.42x10^-3^ |
| PROX2 | 038.2 | Gram positive septicemia | 613 | 19,844 | European | 1.12 | 3.19x10^-3^ |
| PROX2 | 041.1 | *Staphylococcus* infections | 2,180 | 19,844 | European | 1.05 | 2.87x10^-2^ |
| PROX2 | 112 | Candidiasis | 2,284 | 21,426 | European | 0.96 | 4.97x10^-2^ |
| USP44 | 041.2 | *Streptococcus* infection | 1,262 | 19,844 | European | 0.92 | 2.60x10^-3^ |
| USP44 | 038.1 | Gram negative septicemia | 820 | 19,844 | European | 0.92 | 1.83x10^-2^ |
| USP44 | 110 | Dermatophytosis and dermatomycosis | 2,192 | 21,426 | European | 1.05 | 1.84x10^-2^ |
| GPRIN1 | 041.2 | *Streptococcus* infection | 1,262 | 19,844 | European | 1.10 | 1.28x10^-3^ |
| GPRIN1 | 038.1 | Gram negative septicemia | 820 | 19,844 | European | 1.09 | 1.62x10^-2^ |
| NSUN5 | 320 | Meningitis | 114 | 25,170 | European | 0.81 | 2.37x10^-3^ |
| NSUN5 | 041.4 | *Escherichia coli* infection | 1,231 | 19,844 | European | 0.92 | 3.53x10^-3^ |
| NSUN5 | 079 | Viral infection | 1,811 | 20,904 | European | 0.94 | 1.26x10^-2^ |
| NSUN5 | 112 | Candidiasis | 2,284 | 21,426 | European | 0.95 | 1.94x10^-2^ |
| NSUN5 | 054 | Herpes simplex | 610 | 20,904 | European | 0.92 | 2.89x10^-2^ |
| C19orf55/PROSER3 | 117 | Mycoses | 627 | 21,426 | European | 1.10 | 2.77x10^-2^ |
| C19orf55/PROSER3 | 041.4 | *Escherichia coli* infection | 1,231 | 19,844 | European | 1.06 | 4.86x10^-2^ |
| DNAJC17 | 079 | Viral infection | 736 | 3,241 | African | 0.89 | 5.26x10^-3^ |
| DNAJC17 | 481 | Influenza | 125 | 2,991 | African | 0.82 | 3.58x10^-2^ |
| GNRH1 | 079 | Viral infection | 736 | 3,241 | African | 1.13 | 4.53x10^-3^ |
| GNRH1 | 480.1 | Bacterial pneumonia | 112 | 2,991 | African | 0.77 | 5.48x10^-3^ |
| GNRH1 | 078 | Viral warts & HPV | 136 | 3,241 | African | 1.21 | 3.54x10^-2^ |
| LTBP4 | 038.1 | Gram negative septicemia | 820 | 19,844 | European | 1.08 | 3.78x10^-2^ |
| CRYL1 | 112 | Candidiasis | 2,284 | 21,426 | European | 1.05 | 2.39x10^-2^ |
| CRYL1 | 078 | Viral warts and HPV | 1,152 | 20,904 | European | 1.07 | 2.80x10^-2^ |
| LIMK2 | 041.4 | *Escherichia coli* infection | 1,231 | 19,844 | European | 0.91 | 9.46x10^-4^ |
| LIMK2 | 038.1 | Gram negative septicemia | 820 | 19,844 | European | 0.91 | 4.88x10^-3^ |
| LIMK2 | 324 | Other CNS infection and poliomyelitis | 136 | 25,170 | European | 0.82 | 1.44x10^-2^ |
| LIMK2 | 038.1 | Gram negative septicemia | 820 | 19,844 | European | 0.83 | 1.56x10^-2^ |
| LIMK2 | 112 | Candidiasis | 2,284 | 21,426 | European | 0.96 | 4.58x10^-2^ |
| TNNC2 | 070.3 | Viral hepatitis C | 150 | 3,241 | African | 1.34 | 8.39x10^-5^ |
| TNNC2 | 481 | Influenza | 125 | 2,991 | African | 1.34 | 1.64x10^-4^ |
| PRELID2 | 110 | Dermatophytosis and dermatomycosis | 654 | 3,330 | African | 1.13 | 5.04x10^-3^ |
| PRELID2 | 041.4 | *Escherichia coli* infection | 243 | 3,337 | African | 1.19 | 8.98x10^-3^ |
| PRELID2 | 079 | Viral infection | 736 | 3,241 | African | 1.11 | 1.04x10^-2^ |
| PRELID2 | 070.3 | Viral hepatitis C | 150 | 3,241 | African | 1.21 | 1.73x10^-2^ |
| PRELID2 | 041.1 | *Staphylococcus* infections | 358 | 3,337 | African | 1.14 | 2.16x10^-2^ |
| PRELID2 | 041.2 | *Streptococcus* infection | 229 | 3,337 | African | 1.16 | 2.52x10^-2^ |
| PRELID2 | 038.2 | Gram positive septicemia | 114 | 3,337 | African | 1.21 | 4.15x10^-2^ |
| PTCD3 | 041.1 | *Staphylococcus* infections | 2,180 | 19,844 | European | 0.95 | 1.72x10^-3^ |
| PTCD3 | 136 | Other infectious and parasitic diseases | 746 | 24,770 | European | 0.95 | 8.51x10^-3^ |
| PTCD3 | 041.9 | Infection with drug-resistant microorganisms | 893 | 19,844 | European | 0.95 | 1.22x10^-2^ |
| ATG9A | 070.2 | Viral hepatitis B | 166 | 20,904 | European | 0.83 | 2.09x10^-3^ |
| ATG9A | 041.4 | *Escherichia coli* infection | 1,231 | 19,844 | European | 1.08 | 1.28x10^-2^ |
| EIF3M | 110 | Dermatophytosis and dermatomycosis | 654 | 3,330 | African | 1.09 | 4.01x10^-2^ |
| CDK5 | 041.1 | *Staphylococcus* infections | 358 | 3,337 | African | 0.78 | 2.37x10^-5^ |
| CDK5 | 054 | Herpes simplex | 154 | 3,241 | African | 1.33 | 1.22x10^-3^ |
| CDK5 | 136 | Other infectious and parasitic diseases | 173 | 4,054 | African | 0.81 | 5.80x10^-3^ |
| CDK5 | 038.1 | Gram negative septicemia | 139 | 3,337 | African | 0.81 | 1.33x10^-2^ |
| CDK5 | 112 | Candidiasis | 563 | 3,330 | African | 0.90 | 1.96x10^-2^ |
